# Altered m^6^A methylation across eight reward- and motor-related brain regions in alcohol use disorder

**DOI:** 10.64898/2026.09.16.26363234

**Authors:** Abhishek Kumar, Ojong Tabi Ojong Besong, Abhyuday Singh Parihar, Ji Sun Koo, Aravind Panicker, Yohana Kefella, Jennifer E. Beane, Huiping Zhang

## Abstract

N^6^-methyladenosine (m^6^A) is the most abundant internal mRNA modification, dynamically regulating post-transcriptional gene expression. Although m^6^A dysregulation has been implicated in addiction neurobiology, comprehensive characterization of m^6^A alterations in alcohol use disorder (AUD) remains unexplored. We profiled m^6^A methylomes across eight reward- and motor-related brain regions in postmortem tissue from 12 AUD subjects and 12 matched controls (192 samples total) using MazF-mediated enzymatic cleavage and microarray-based m^6^A site mapping. Region-specific differential analysis identified 1,403 differentially methylated m^6^A sites (*P* < 0.05, |fold change| ≥ 1.5): amygdala (n = 198), caudate nucleus (n = 91), cerebellum (n = 201), hippocampus (n = 89), nucleus accumbens (n = 276), prefrontal cortex (n = 362), putamen (n = 142), and ventral tegmental area (n = 241). Host transcripts were enriched for functional categories spanning intercellular communication, immune and inflammatory responses, RNA and protein synthesis, energy metabolism, and neurodegeneration. Gene set enrichment analysis revealed a coordinated inverse relationship between m^6^A methylation status and transcript abundance: hypermethylated transcripts were enriched among downregulated genes and hypomethylated transcripts among upregulated genes, consistent with m^6^A-mediated transcript destabilization. Cross-validation with Allen Human Brain Atlas region-specific expression profiles demonstrated that differential methylation patterns were primarily AUD-driven rather than secondary to regional transcriptional heterogeneity. These findings position m^6^A epitranscriptomics as a novel molecular signature of AUD and suggest that m^6^A regulatory machinery may serve as a therapeutic target. Future studies with larger validation cohorts and mechanistic interrogation are essential to establish causal relationships and facilitate translation to drug development.

## INTRODUCTION

Alcohol use disorder (AUD) emerges from complex interactions between genetic predisposition and environmental exposures. In addition to inherited variation, repeated alcohol consumption and environmental stressors produce persistent epigenetic changes that alter gene regulation at multiple levels, including DNA methylation, histone modifications, noncoding RNA dysregulation, and RNA chemical modifications, and thereby contribute to the neurobiological adaptations underlying addiction. To date, most epigenetic investigations in AUD have focused on DNA methylation and histone marks. Genome-wide DNA methylation studies have profiled peripheral blood ^1, 2^, saliva ^3^, and postmortem brain ^4–7^, revealing locus specific and pathway level differences between AUD cases and controls. Alcohol induced alterations in histone methylation and acetylation have likewise been reported across animal models and human tissues, with downstream effects on transcriptional programs linked to addiction relevant behaviors ^8–11^. Emerging work also implicates noncoding RNAs, including microRNAs (miRNAs) and long noncoding RNAs (lncRNAs), in AUD across saliva and brain studies ^12–19^. By contrast, the epitranscriptome (chemical modifications on RNA) remains comparatively underexplored in human AUD.

N^6^ methyladenosine (m^6^A) is the most abundant internal modification of eukaryotic mRNA and a key regulator of post transcriptional fate, influencing mRNA stability, splicing, translation, and subcellular localization. m^6^A sites are typically enriched near stop codons and in 3′ untranslated regions (3’ UTRs), often occur within the DRA*CH consensus motif (where A* is the modified adenosine, D=A/G/U, R=A/G, H=A/C/U), and have well documented roles in development, synaptic function, and stress responses ^20–24^. Transcriptome wide m^6^A mapping is now feasible *via* antibody based immunoprecipitation (meRIP seq/m^6^A seq, m^6^A CLIP), enzyme based cleavage assays (e.g., MAZTER seq), and direct RNA nanopore sequencing ; each approach carries tradeoffs in resolution, input requirements, and specificity ^25–29^.

Only in recent years has research begun to investigate the relationship between m^6^A RNA methylation and substance use disorders (SUDs). Animal and cellular studies indicate that psychostimulant and alcohol exposures remodel m^6^A landscapes in brain regions implicated in reward and plasticity. For example, m^6^A profiling in cocaine conditioned mice revealed altered peaks in genes controlling synaptic localization and maturation ^30^. In human and cellular models of alcohol exposure, preliminary reports suggest m^6^A alterations at receptors and immune related transcripts: our prior pilot analysis of human nucleus accumbens (NAc) identified AUD associated m^6^A changes enriched for immune signaling pathways ^31^, and chronic intermittent ethanol (CIE) exposure in cultured cells increased global m^6^A levels while producing localized hypomethylation near opioid receptor transcripts ^32^. These findings collectively implicate m^6^A dysregulation in substance induced reprogramming of reward related gene networks, but human data are limited by small sample sizes and single region designs.

Multiple interconnected reward and motor related brain regions mediate distinct components of alcohol related behavior, and region specific epigenetic remodeling may underlie the heterogeneous cognitive, emotional, and motor sequelae of AUD. The amygdala (AMY) contributes to stress and negative affect driven drinking and relapse; the hippocampus (HIP) encodes contextual memories that support cue triggered craving; the ventral tegmental area (VTA) and nucleus accumbens (NAc) form a core mesolimbic circuit for dopamine mediated reinforcement and motivation; the prefrontal cortex (PFC) governs executive control and inhibitory processes that are compromised in AUD; and dorsal striatal regions [caudate nucleus (CN) and putamen (PUT)] and the cerebellum (CRB) support habit formation, sensorimotor integration, and alcohol sensitive motor control. Profiling m^6^A across these regions in human AUD cases and matched controls therefore offers an opportunity to link epitranscriptomic variation to region specific molecular pathways and to transcriptional dysregulation implicated in addiction.

We performed a systematic, multi region epitranscriptomic survey of postmortem human brains from individuals with AUD and matched controls. Using MazF-mediated digestion coupled with microarray-based m^6^A site mapping, we systematically profiled region-specific m^6^A modifications across eight functionally distinct brain regions (reward and motor circuits) in individuals with AUD. Differentially methylated transcripts converged on intercellular communication, neuroinflammatory responses, RNA and protein synthesis, and metabolic signaling. Critically, m^6^A methylation gains and losses were consistently correlated with changes in transcript abundance, indicating that altered m^6^A patterns represent AUD-associated post-transcriptional regulatory mechanisms. These results implicate regionally patterned m^6^A remodeling as an underappreciated layer of molecular plasticity contributing to AUD pathophysiology.

## MATERIALS AND METHODS

### Human postmortem tissue

Postmortem brain tissue samples were procured from the New South Wales Brain Tissue Resource Centre (NSWBTRC), a biorepository at the University of Sydney, Australia. The cohort comprised 24 donors: 12 individuals with alcohol use disorder (AUD) and 12 matched controls (all Caucasian Australians; AUD: 6 males, 6 females; controls: 6 males, 6 females). For each donor, tissue was dissected from eight brain regions (AMY, CN, CRB, HIP, NAc, PFC, PUT, and VTA) implicated in reward and motor function, yielding 192 samples (8 regions × 24 donors). Controls were matched to cases by age, sex, and ethnicity and had no history of AUD. Donor metadata [daily alcohol consumption, sex, age, postmortem interval (PMI), RNA integrity number (RIN), brain weight, brain pH, sampled hemisphere, smoking status, liver disease] are provided in Supplementary Table 1. Psychiatric and substance use histories were ascertained per NSWBTRC protocols and DSM IV criteria ^33^.

### Total RNA extraction and quality assessment

Total RNA was isolated from 10-50 mg of frozen tissue per sample (n = 192) using the miRNeasy Mini Kit (QIAGEN, Valencia, CA, USA) according to the manufacturer’s protocol. RNA concentrations and purity were measured on a NanoDrop ND 1000 spectrophotometer (Thermo Fisher Scientific, Waltham, MA, USA); purity was evaluated using the OD_260nm_/OD_280nm_ ratio. RNA integrity was assessed with an Agilent 2100 Bioanalyzer and the RNA 6000 Nano Kit (Agilent Technologies, Santa Clara, CA, USA). Across the cohort, mean OD_260nm_/OD_280nm_ was 1.99 ± 0.10 (mean ± SD) and mean RNA integrity number (RIN) was 4.8 ± 1.4 (mean ± SD). Aliquots of total RNA were prepared and stored at −80°C until downstream assays.

### Global m^6^A quantification

Global m^6^A content was quantified with the EpiQuik m^6^A RNA Methylation Quantification Kit (EpigenTek, Farmingdale, NY, USA) following the manufacturer’s instructions. Briefly, input RNA was immobilized in 96 well plates, followed by incubation with an m^6^A specific capture antibody and detection reagents. Color development was quantified by measuring absorbance at 450 nm on a SpectraMax i3 microplate reader (Molecular Devices, San Jose, CA, USA). Each plate included positive and negative controls from the kit. A standard curve was generated from serial dilutions of the positive control and modeled using a four parameter logistic (4PL) regression [y = (0.046794 − 1.518579) / (1 + (x/ 0.115722)^ 3.584020) + 1.518579] implemented in Arigo’s ELISA Calculator; the 4PL model provided the best fit (Supplementary Figure S1). Sample m^6^A amounts (ng) were calculated from OD_450nm_ values after subtracting the negative control reading and normalized to input RNA. Percent m^6^A was computed using the formula: m^6^A% = [m^6^A amount (ng) / S (ng)] × 100, where S denotes the input RNA amount. For each brain region, group comparisons of global m^6^A levels between AUD and control subjects (n = 12 per group) were performed using two sided Wilcoxon rank-sum tests (Mann-Whitney U) implemented in R (wilcox.test() function, R version 4.4.0). For each region, the test statistic (W), Hodges-Lehmann location shift estimate, and corresponding two sided *P* values were reported. *P* values were adjusted across the eight brain regions using the Benjamini-Hochberg false discovery rate (FDR) ^34^.

### m^6^A epitranscriptome microarray profiling and raw data processing

Because RNA from archived postmortem tissue was partially degraded, we profiled the m^6^A epitranscriptome using the Arraystar Human m^6^A Single Nucleotide Array (Arraystar, Rockville, MD, USA). This assay leverages the methylation sensitive endoribonuclease MazF to detect and quantify m^6^A at ACA motifs, and it does not require antibody immunoprecipitation, making it tolerant of RNA fragmentation. The array contains 14,321 distinct probes (60 nt each) targeting 11,237 single ACA sites (> 40 nt separation between adjacent sites) and 3,084 poly ACA sites (multiple ACA sites within a 20 nt window), collectively covering 5,494 mRNAs and 164 noncoding RNAs (ncRNAs). Probe annotations were derived from experimentally validated m^6^A sites identified in published miCLIP studies ^21, 35, 36^ and the RMBase database ^37, 38^.

For each sample, 2 μg of total RNA was split into two aliquots (1 μg each). One aliquot was treated with MazF endoribonuclease (MazF digested) to cleave unmethylated ACA sites, while the other remained untreated (MazF undigested). Both aliquots were spiked with equal amounts of calibration spike-in control RNA, then amplified and fluorescently labeled using the Arraystar Super RNA Labeling Kit (Arraystar): MazF-digested cRNA with Cy5 and MazF-undigested cRNA with Cy3. Paired Cy5/Cy3 cRNAs pools were co hybridized onto the array slide at 60°C for 17 hours in an Agilent hybridization oven (Agilent Technologies, Santa Clara, CA, USA), followed by washing and two color scanning on an Agilent G2505C scanner (Agilent Technologies, Santa Clara, CA, USA). Array images were acquired using the Agilent Feature Extraction software (version 11.0.1.1).

Raw signal intensities of MazF-digested (Cy5) and MazF-undigested (Cy3) channels were normalized to the mean log_2_-scaled Spike-in RNA intensities. Following spike-in normalization, probes meeting a predefined proportion threshold for Present (P) or Marginal (M) quality control (QC) flags were retained for downstream analysis. m^6^A methylation stoichiometry at each ACA site was quantified as the m^6^A percentage (%m^6^A), defined as: %m^6^A = (Cy5/Cy3) × correction factor, where the per-sample correction factor is the ratio of MazF-undigested to MazF-digested input amount. Because this correction factor can exceed unity, %m^6^A values may exceed 100% at highly methylated sites. Unlike Cy5 intensity alone, which confounds methylation status with transcript abundance, %m6A isolates the methylated fraction at each site independently of expression level.

Quality control (QC), probe filtering, and batch correction of the 192 samples were performed as follows. Samples were processed in two technical batches stratified by brain regions: Batch 1 (AMY, HIP, PFC, and VTA; n = 4 × 24 = 96 samples) and Batch 2 (CN, CRB, NAc, and OUT; n = 4 × 24 = 96 samples). Probe level QC included inspection of intensity distributions and missingness, followed by a two-step filter applied uniformly across all samples: (1) removal of zero-variance probes and (2) exclusion of the lowest 10% of probes ranked by interquartile range (IQR). Principal component analysis (PCA) of the filtered probe matrix was performed to assess technical variation prior to batch correction. Batch effects were removed using ComBat from the sva package (sva v3.52.0) ^39^, with AUD diagnosis specified as a protected covariate to preserve biological variation of interest. PCA was repeated following ComBat to confirm successful reduction of batch associated variance (Supplementary Figure 2). For downstream linear modeling, per-probe %m^6^A values were log_2_-transformed and quantile-normalized within each brain region across the 24 samples. Shapiro-Wilk tests confirmed that the resulting log_2_(%m6A) values were approximately normally distributed, satisfying linear model assumptions.

### Statistical analysis of differentially methylated m^6^A sites

Following QC, 10,249 probes common to all eight brain regions were retained by inner join: 10,007 targeting m^6^A sites in 4,489 coding mRNAs and 242 targeting m^6^A sites in 124 ncRNAs. Among the 10,007 mRNA-targeting probes, 283 (2.8%) mapped to 5’ untranslated regions (5’ UTRs), 5,837 (58.3%) to coding sequences (CDS), and 3,887 (38.8%) to 3′ untranslated regions (3’ UTRs). Differential m^6^A methylation between AUD subjects and controls was assessed separately for each brain region using linear modeling implemented in limma (3.60.0, Bioconductor) ^40^. To accommodate the repeated-measures structure of the data, i.e., each donor contributing tissue from all eight brain regions, subject identity was specified as a blocking factor, and within-donor correlation was estimated with duplicateCorrelation(). A design matrix encoding region-by-diagnosis interaction terms (e.g., amygdala.AUD, amygdala.Control) was constructed to permit direct within-region AUD-versus-control contrasts. Sex and RNA integrity number (RIN) were included as fixed-effect covariates to control for potential demographic and tissue-quality confounding, respectively. Models were fitted with lmFit() incorporating both the blocking structure and the estimated within-donor correlation. Region-specific contrasts were defined using makeContrasts() and applied *via* contrasts.fit(). Empirical Bayes moderation of variance estimates was performed with eBayes(), and differential methylation was evaluated using moderated t-statistics. *P* values were corrected for multiple comparisons using the Benjamini-Hochberg FDR procedure ^34^. All design matrices, contrast specifications, and probe-filtering criteria were pre-specified prior to analysis to ensure reproducibility. m^6^A sites with nominal *P* < 0.05 and |fold change| ≥ 1.5 were considered differentially methylated.

### m^6^A transcript level aggregation and pathway enrichment analysis

To facilitate biological interpretation, probe level differential methylation results were aggregated to the transcript level using Arraystar probe-to-transcript annotations. For transcripts represented by multiple m^6^A probes, the probe with the largest absolute moderated t-statistic (|t|) was selected as the representative feature; its sign was preserved to reflect directionality (hypermethylation: positive; hypomethylation: negative), thereby avoiding transcript-level redundancy in downstream analysis. The resulting per-transcript signed moderated t-statistics were compiled into region-specific pre-ranked gene lists, one per brain region, for subsequent gene set enrichment analysis (GSEA). GSEA was performed using the fgsea package (v1.30.0) ^41^ with pre-ranked transcript lists as input. Pathway gene sets were drawn from curated collections including the Kyoto Encyclopedia of Genes and Genomes (KEGG) database. Gene sets were restricted to those containing between 15 and 500 genes (minSize = 15; maxSize = 500) to exclude overly small or broad gene sets that may produce unreliable enrichment estimates. Normalized enrichment scores (NES) were computed to account for differences in gene set size, and *P* values were adjusted for multiple comparisons using the Benjamini-Hochberg FDR procedure. Enrichment curves for selected pathways were visualized using fgsea’s plotEnrichment() function. To provide an integrated visual summary, a bubble plot was constructed depicting enriched pathways (*P* < 0.05) identified among differentially methylated transcripts across all eight brain regions, with bubble size and color reflecting gene set size and normalized enrichment score (NES), respectively.

### Analysis of regional enrichment of differentially methylated transcripts among differentially expressed genes

To examine whether region specific m^6^A methylation changes are associated with region specific mRNA expression alterations in AUD, we performed GSEA integrating differential methylation results from the present study with differential expression rankings from our prior RNA-seq study ^19^. For each brain region, all genes analyzed in the differential expression pipeline were ranked in descending order by their moderated t-statistics, yielding region-specific pre-ranked gene lists (gene counts: amygdala, 14,727; caudate nucleus, 14,730; cerebellum, 14,726; hippocampus, 14,729; nucleus accumbens, 14,730; prefrontal cortex, 14,724; putamen, 14,730; ventral tegmental area, 14,729). For transcripts represented by multiple m^6^A probes in the methylation dataset, the probe with the largest absolute moderated t-statistic (|t|) was selected as the representative feature, as described above. Query gene sets for GSEA comprised transcripts with nominally significant differential m^6^A methylation (*P* < 0.05, unadjusted) in each brain region. To distinguish directional effects, hypermethylated (*P* < 0.05, t > 0) and hypomethylated (*P* < 0.05, t < 0) transcript subsets were tested separately against the corresponding regional expression rankings. This directional approach was motivated by the canonical role of m^6^A in promoting mRNA decay. Normalized enrichment scores (NES) and Benjamini-Hochberg-adjusted FDRs were reported for all enrichment tests.

### Enrichment of differentially methylated transcripts in region-preferentially expressed genes: Allen Human Brain Atlas analysis

To determine whether AUD-associated m^6^A methylation alterations reflect AUD-driven epitranscriptomic changes rather than inherent regional gene expression patterns, we performed an integrative GSEA using region-level differential expression data from the Allen Human Brain Atlas (AHBA; https://human.brain-map.org/). For each of the eight brain regions, differential expression results were retrieved using the following AHBA query parameters: Target Structure = [region name], Contrast Structure = [whole brain], and Selected Donors = [all donors]. This approach yielded region-preferential gene rankings of varying sizes: amygdala (AMY) (18,405 genes), caudate nucleus (CN) (14,148 genes), cerebellum (CRB) (15,669 genes), hippocampus (HIP) (14,804 genes), nucleus accumbens (NAc) (15,296 genes), prefrontal cortex (PFC)/frontal lobe (15,443 genes), putamen (PUT) (15,630 genes), and ventral tegmental area (VTA) (21,043 genes). Because AHBA differential expression results do not include moderated t-statistics, region-specific gene lists were ranked by ascending *P* value (smallest to largest) to prioritize genes with the strongest evidence of region-preferential expression. Transcripts with nominally significant differential m^6^A methylation (*P* < 0.05, unadjusted) in each brain region were assembled as query gene sets and tested for enrichment among region-preferentially expressed genes using fgsea-based GSEA, as described above. Enrichment results were reported as normalized enrichment scores (NES) with Benjamini-Hochberg-adjusted FDRs, and enrichment curves were visualized using fgsea’s plotEnrichment() function. Significant enrichment of differentially methylated transcripts within a given region’s preferentially expressed gene set would suggest that the observed m^6^A alterations track regional transcriptional identity rather than AUD-specific epitranscriptomic dysregulation; conversely, absence of enrichment would support an AUD-driven interpretation.

## RESULTS

### Global m^6^A RNA methylation in AUD brains

Global m^6^A levels were quantified by an ELISA-like assay and compared between AUD cases and controls across eight reward- and motor-related brain regions using Wilcoxon rank-sun tests. No significant differences in global m^6^A levels were detected between AUD cases and controls in any region (Supplementary Figure 3). The hippocampus (HIP) showed a trend toward elevated global m^6^A levels in AUD subjects (median = 0.049) compared with controls (median = 0.039), though this did not reach statistical significance (*P* = 0.053).

### Differentially methylated m^6^A sites across eight brain regions of AUD subjects

Although global m^6^A methylation levels did not differ significantly between AUD cases and controls, we identified AUD-associated differential m^6^A methylation at specific sites of the transcriptome across all eight brain regions. Volcano plots display results for 10,249 m^6^A sites (10,007 mapping to 4,489 mRNAs and 242 mapping to 124 ncRNAs), revealing region-specific patterns of differential methylation (Figure 1). Applying thresholds of unadjusted *P* < 0.05 and |fold change| ≥ 1.5, the numbers of AUD-associated differentially methylated m^6^A sites per region were: amygdala (AMY), 198 sites (61 hyper-, 137 hypomethylated); caudate nucleus (CN), 91 (22/69); cerebellum (CRB), 201 (140/61); hippocampus (HIP), 89 (33/56); nucleus accumbens (NAc), 276 (119/157); prefrontal cortex (PFC), 362 (205/157); putamen (PUT), 142 (87/55); and ventral tegmental area (VTA), 241 (155/86). In total, 1,403 unique differentially methylated m^6^A sites were identified across the eight brain regions. The 10 most significantly dysregulated m^6^A sites (|fold change| ≥ 1.5 and with the smallest *P*-values) in each brain region are listed in Supplementary Table 2.

**Fig. 1.**
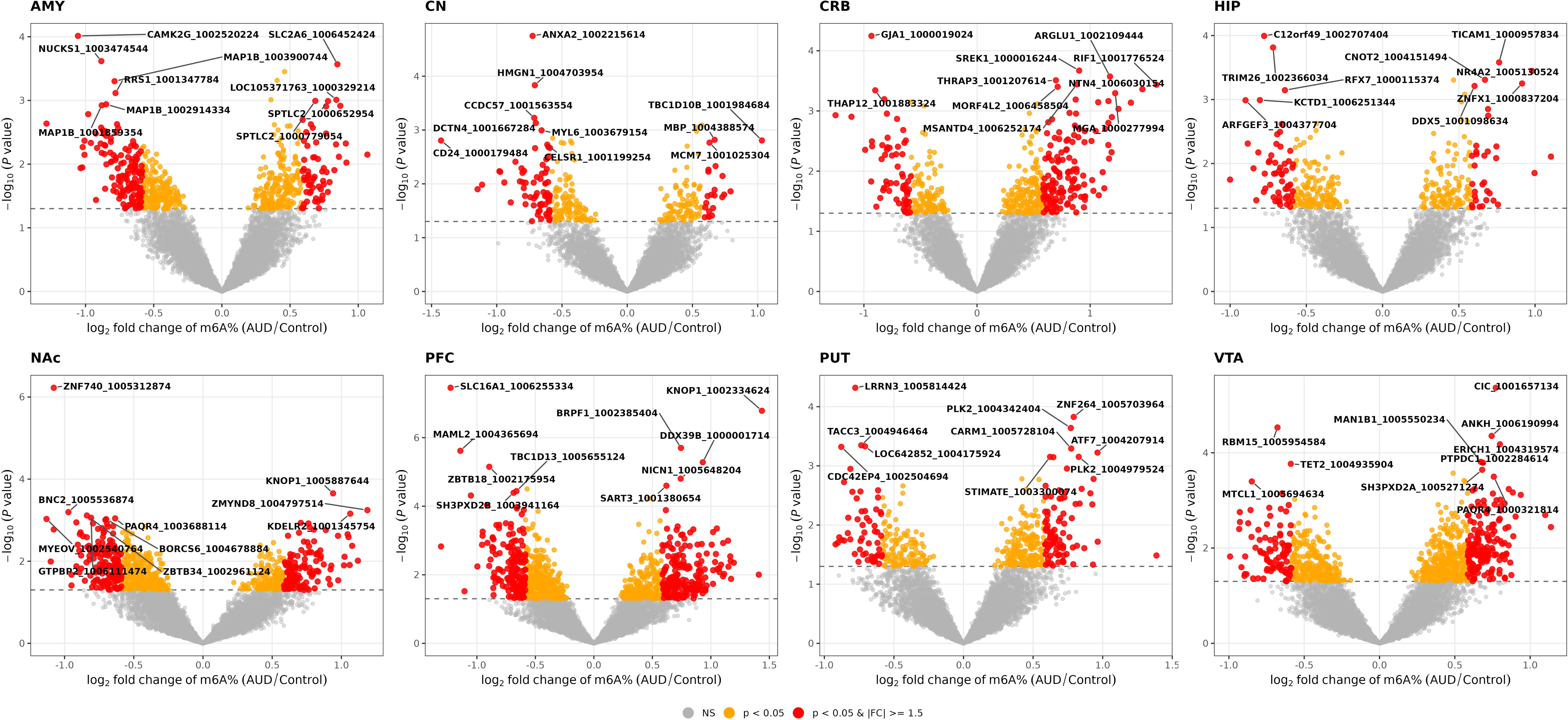
Volcano plots of regional m^6^A methylation changes in AUD across eight brain regions. Each point represents a m^6^A site. The x-axis shows log_2_ fold change (log_2_FC; AUD vs. control), and the y-axis shows −log_10_(*P*-value). Color coding reflects significance and effect size: red, *P* < 0.05 and |FC| ≥ 1.5 (strongly dysregulated); orange, *P* < 0.05 and |FC| < 1.5 (nominally significant); gray, *P* ≥ 0.05 (non-significant). The probe names for the top 10 differentially methylated sites are labeled. Panels correspond to eight brain regions: amygdala (AMY), caudate nucleus (CN), cerebellum (CRB), hippocampus (HIP), nucleus accumbens (NAc), prefrontal cortex (PFC), putamen (PUT), and ventral tegmental area (VTA).

The counts of hyper- and hypomethylated sites (unadjusted *P* < 0.05, |fold change| ≥ 1.5) in each brain region are shown in Supplementary Figure 4. Among the eight regions, the PFC had the largest number of hypermethylated sites (n = 205), while the CN had the fewest (n = 22). Both the PFC and NAc exhibited the largest number of hypomethylated sites (n = 157 each), whereas the PUT had the fewest (n = 55). The counts of transcripts with exclusively hyper-, exclusively hypo-, or both hyper- and hypomethylated sites are shown in Supplementary Figure 5. Most transcripts harbored exclusively hyper- or hypomethylated sites. Transcripts containing both hyper- and hypomethylated sites were rare, with two identified in the AMY, five in the PFC, and one each in the HIP, NAc, PUT, and VTA. Additionally, the distributions of differentially methylated m^6^A sites across the eight brain regions are illustrated by chromosome circos diagrams, with rings ordered from outermost to innermost as follows: chromosomal location, transcript names and positions, hyper- or hypomethylated sites, *P* values, and fold changes (Supplementary Figure 6).

To characterize overlapping versus distinct patterns of m^6^A methylation dysregulation across the eight brain regions, an UpSet plot was generated (Figure 2). Differential methylation was predominantly region-specific: of the 1,403 differentially methylated m^6^A sites (unadjusted *P* < 0.05, |fold change| ≥ 1.5), 1,214 (86.5%) were unique to a single region (region-specific fractions ranging from 65.9% in the CN to 81.8% in the PFC), while 189 sites were shared across two or three regions. Among the shared sites, eight (mapping to transcripts of *CA12*, *IRS2*, *MORC2*, *MTR*, *MYEOV*, *NTN4*, *RAPGEF1*, and *XIST*) were differentially methylated in three regions, and the remaining 181 were shared by two regions. No sites were shared in more than three regions. This predominance of region-specific dysregulation indicates that AUD-associated m^6^A changes are largely region-restricted, with the direction of methylation varying by region.

**Fig. 2.**
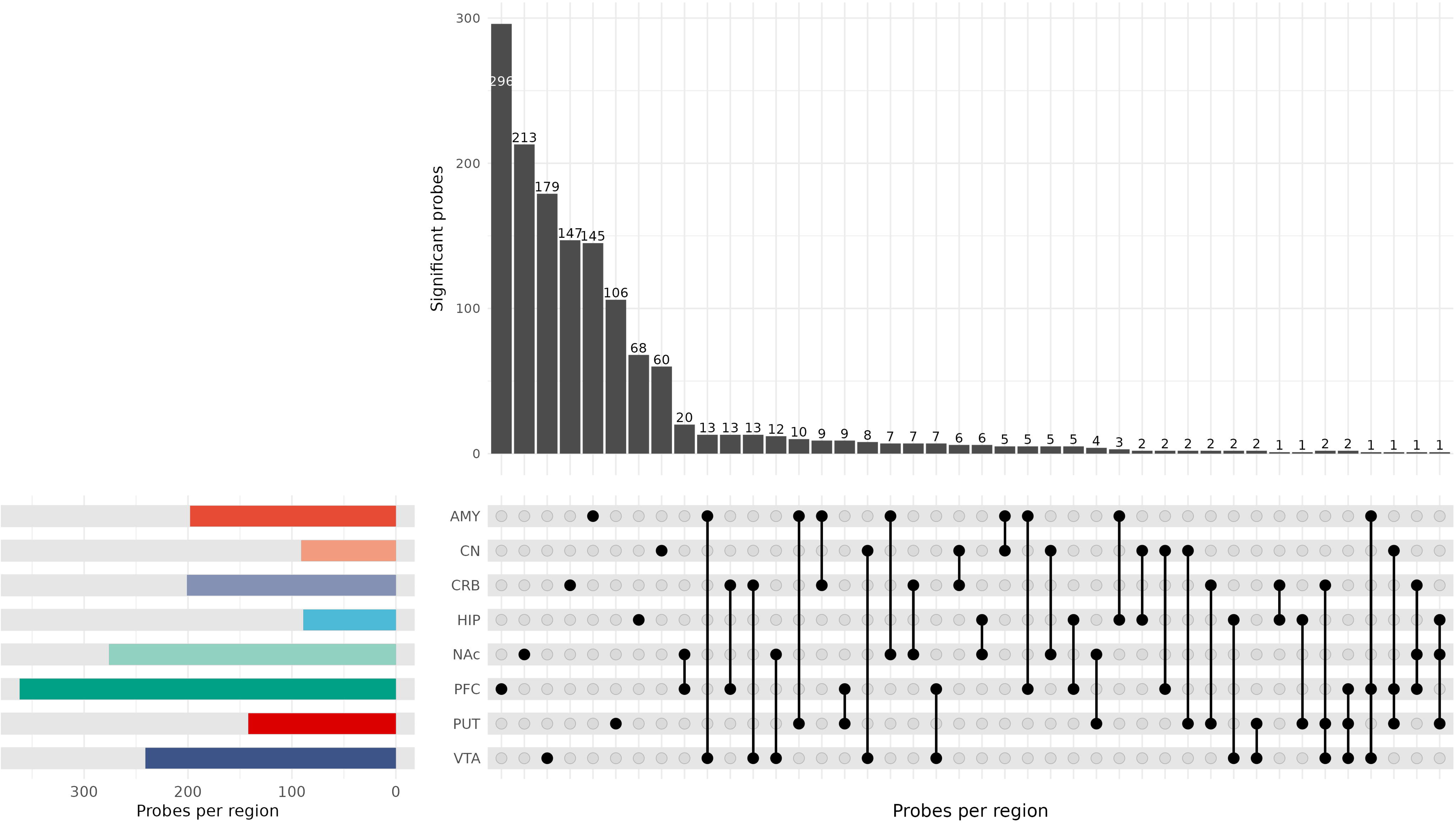
UpSet plot illustrating shared and region-specific patterns of m^6^A methylation dysregulation across eight brain regions in AUD. Only differentially methylated sites meeting both significance (*P* < 0.05) and effect size (|FC| ≥ 1.5) thresholds are included. Vertical bars indicate the number of dysregulated m^6^A sites unique to or shared among brain regions. Horizontal bars on the left show the total number of dysregulated sites per region. Brain regions examined: amygdala (AMY), caudate nucleus (CN), cerebellum (CRB), hippocampus (HIP), nucleus accumbens (NAc), prefrontal cortex (PFC), putamen (PUT), and ventral tegmental area (VTA).

### Functional enrichment of m^6^A-dysregulated transcripts in AUD

Differentially methylated m^6^A sites were mapped to cognate mRNA transcripts and subjected to GSEA against KEGG pathways to identify enriched biological processes (Figure 3). Regional enrichment patterns revealed marked heterogeneity across the eight brain regions. The amygdala (AMY) showed enrichment for *ADRB3-UCP1 Signaling*, *Dilated Cardiomyopathy*, *Gap Junction*, *Ribosome*, *RIG-I-like Receptor Signaling*, *TGF-*β *signaling*, and *Translation Initiation*. The caudate nucleus (CN) was enriched for *ErbB Signaling*, *GF-RTK-RAS-ERK Signaling*, *GF-RTK-RAS-PI3K Signaling*, and *ECM receptor Interaction*. The cerebellum (CRB) was enriched for *Adherens Junction* and *Adipocytokine Signaling*. The hippocampus (HIP) was enriched for *Antigen Processing and Presentation*, *Focal Adhesion*, and *Histone H2AK119 Deubiquitination*. The nucleus accumbens (NAc) showed enrichment for *ADRB3-UCP1 Signaling*, *Dilated Cardiomyopathy*, *Leukocyte Transendothelial Migration*, *Melanogenesis*, *Parkinson’s Disease*, *Spliceosome*, *Ubiquitin-mediated Proteolysis*, *Variant LRP6 Overexpression to Wnt Signaling*, and *Wnt Signaling*. The prefrontal cortex (PFC) was enriched for *Adipocytokine Signaling*, *NK Cell-mediated Cytotoxicity*, *TGF-*β *Signaling*, and *Wnt Signaling*. The putamen (PUT) displayed enrichment for *Focal Adhesion*, *GF-RTK-RAS-ERK Signaling*, *Huntington’s Disease*, *Leukocyte Transendothelial Migration*, *Actin Cytoskeleton Regulation*, and *Vasopressin-Regulated Water Reabsorption*. The ventral tegmental area (VTA) was enriched for *Glycerophospholipid Metabolism*.

**Fig. 3.**
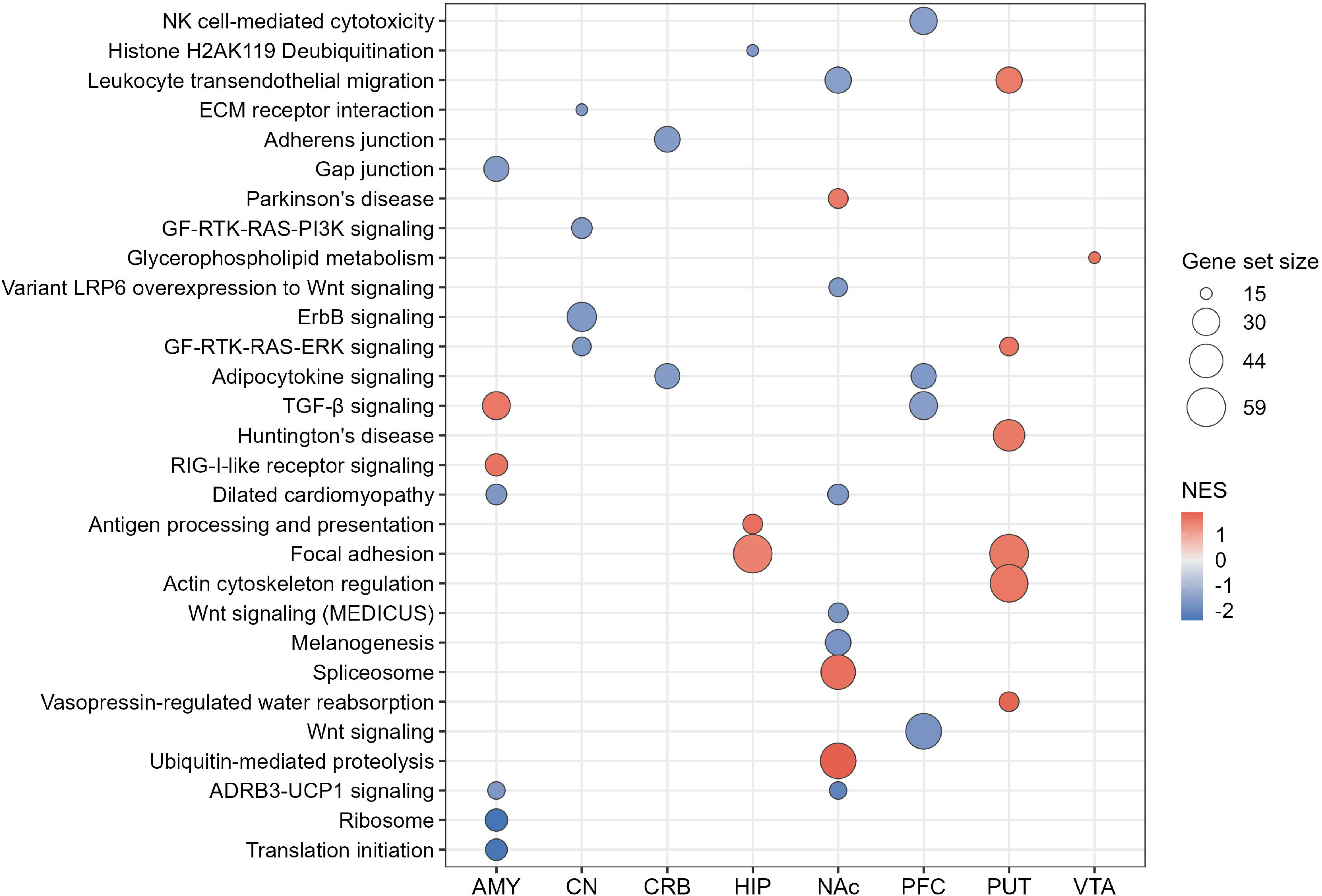
KEGG pathway enrichment of differentially methylated transcripts across eight brain regions in AUD. Each bubble represents a significantly enriched KEGG pathway (*P* < 0.05). Bubble size reflects the number of leading-edge genes contributing to the enrichment signal. Bubble color indicates the normalized enrichment score (NES), with warm colors (positive NES) representing pathways enriched among hypermethylated transcripts and cool colors (negative NES) representing pathways enriched among hypomethylated transcripts. Brain region abbreviations: AMY, amygdala; CN, caudate nucleus; CRB, cerebellum; HIP, hippocampus; NAc, nucleus accumbens; PFC, prefrontal cortex; PUT, putamen; VTA, ventral tegmental area.

Despite this region-specific heterogeneity, the enriched pathways converged on five broad functional systems: (1) cell adhesion and intercellular communication (*Adherens Junction*, *Gap Junction*, *Focal Adhesion*, and *ECM-receptor Interaction*); (2) growth-factor and morphogen signaling (*ErbB*, *RTK-RAS-ERK*, *RTK-RAS-PI3K*, *TGF-*β, and *Wnt Signaling*); (3) immune and inflammatory signaling (*RIG-I-like Receptor*, *Antigen Processing and Presentation*, *NK Cell-mediated Cytotoxicity*, and *Adipocytokine Signaling*); (4) RNA processing and protein homeostasis (*Ribosome*, *Translation Initiation*, *Spliceosome*, *Ubiquitin-mediated Proteolysis*, and *Histone H2AK119 Deubiquitination*); and (5) energy metabolism and neurodegeneration-associated pathways (*ADRB3-UCP1 Signaling*, *Vasopressin-regulated Water Reabsorption*, *Parkinson’s Disease*, and *Huntington’s Disease*). Collectively, these findings indicate that AUD-associated m^6^A dysregulation, while region-specific in its individual targets, converges on a limited set of interconnected molecular programs governing intercellular communication, growth factor and immune signaling, gene expression regulation, and cellular metabolism.

### m^6^A modification-gene expression correlation in AUD brain

To assess whether AUD-associated m^6^A methylation changes were concordant with differential gene expression, we performed GSEA across all eight brain regions. Regional results revealed a consistent reciprocal pattern: hypermethylated transcripts were preferentially enriched among downregulated genes (Figure 4), while hypomethylated transcripts were enriched among upregulated genes (Figure 5), consistent with the canonical role of m^6^A promoting mRNA decay.

**Fig. 4.**
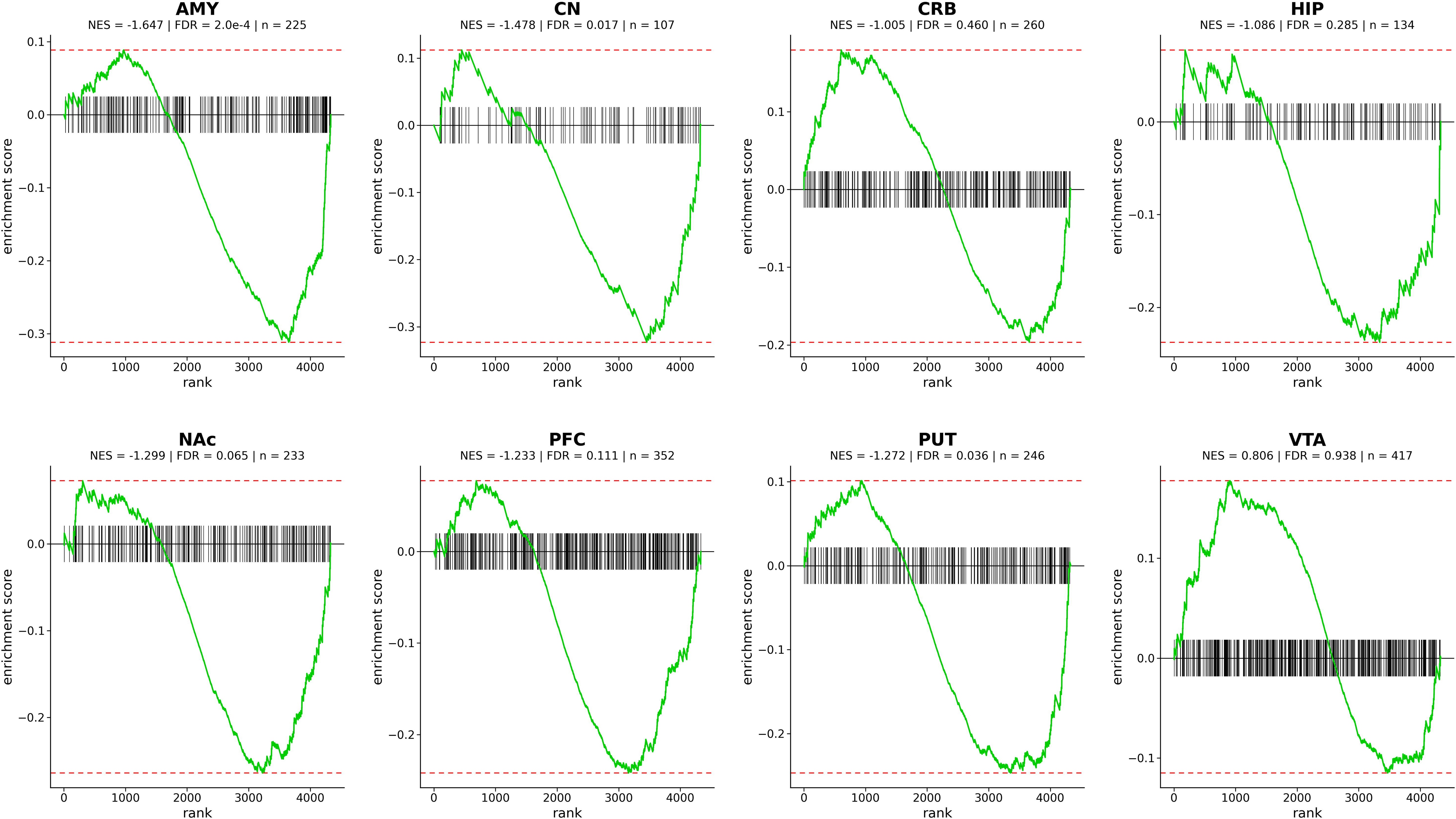
Enrichment of hypermethylated transcripts among differentially expressed genes across eight brain regions in AUD. Each panel displays the GSEA enrichment curve for one brain region, with the running enrichment score plotted along the y-axis and the ranked gene list along the x-axis. Negative normalized enrichment scores (NES < 0) indicate that hypermethylated transcripts are preferentially enriched among downregulated genes in AUD. Vertical tick marks indicate the positions of hypermethylated transcripts within the ranked list. Brain region abbreviations: AMY, amygdala; CN, caudate nucleus; CRB, cerebellum; HIP, hippocampus; NAc, nucleus accumbens; PFC, prefrontal cortex; PUT, putamen; VTA, ventral tegmental area.

**Fig. 5.**
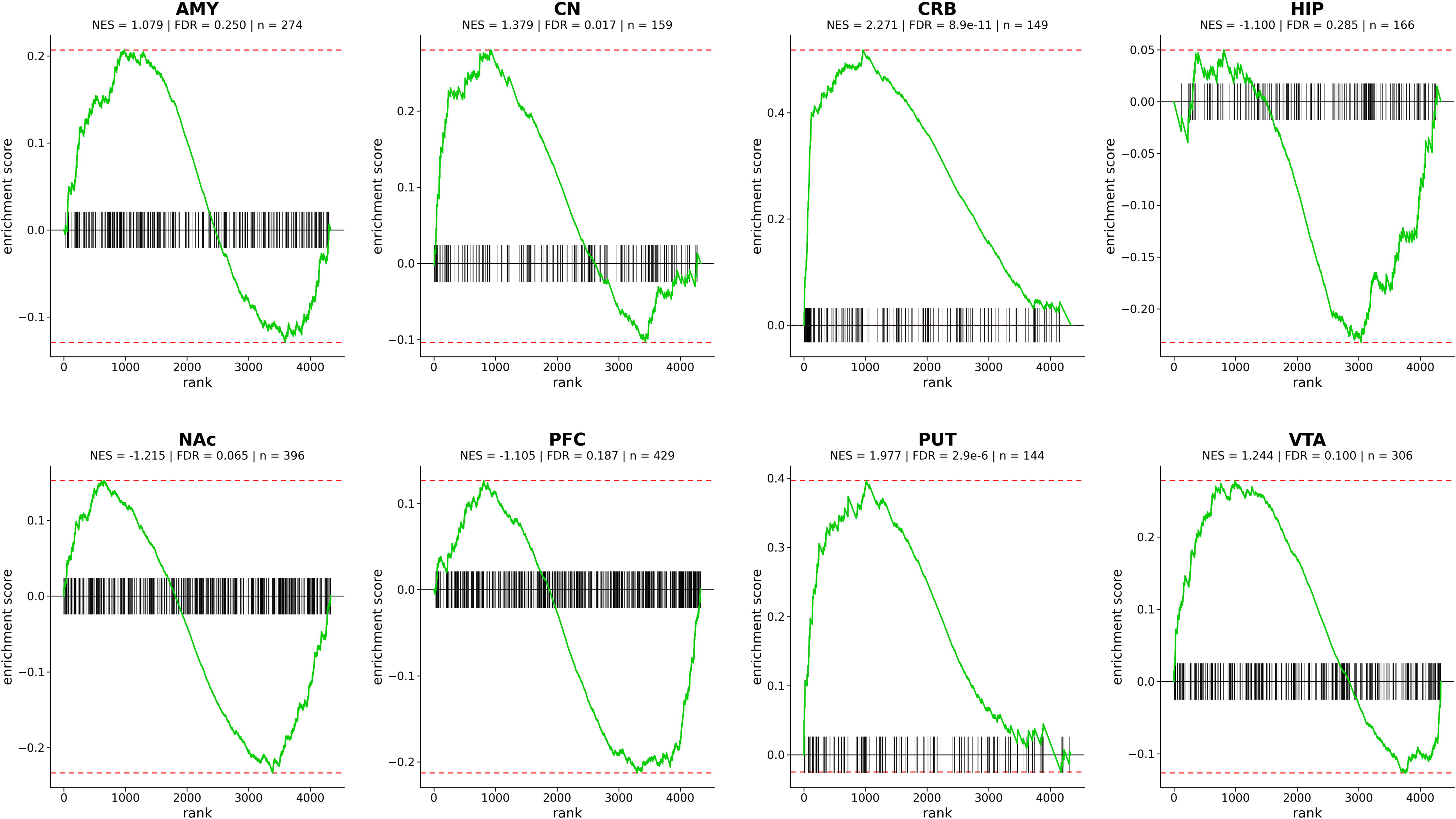
Enrichment of hypomethylated transcripts among differentially expressed genes across eight brain regions in AUD. Each panel displays the running enrichment score curve for one brain region. Positive normalized enrichment scores (NES > 0) indicate that hypomethylated transcripts are preferentially enriched among upregulated genes in AUD. Vertical tick marks denote the positions of individual hypomethylated transcripts within the ranked gene list. Brain region abbreviations: AMY, amygdala; CN, caudate nucleus; CRB, cerebellum; HIP, hippocampus; NAc, nucleus accumbens; PFC, prefrontal cortex; PUT, putamen; VTA, ventral tegmental area.

Specifically, hypermethylation-with-downregulation was significant in the amygdala (AMY) (NES = −1.65, FDR = 2.0×10^-4^), the caudate nucleus (CN) (NES = −1.48, FDR = 0.017), and the putamen (PUT) (NES = −1.27, FDR = 0.036) (Figure 4). Hypomethylation-with-upregulation was significant in the caudate nucleus (CN) (NES = 1.38, FDR = 0.017), the cerebellum (CRB) (NES = 2.27, FDR = 8.9×10^-11^), and the putamen (PUT) (NES = 2.0, FDR = 2.9×10^-6^) (Figure 5). Together, these findings demonstrate that AUD-associated m^6^A dysregulation is mechanistically coupled to altered gene expression through the canonical m^6^A-mediated mRNA decay axis.

### No enrichment of differentially methylated transcripts in region-preferentially expressed genes

To determine whether AUD-associated m^6^A methylation alterations could be attributed to inherent regional gene expression patterns, we performed GSEA using region-preferential expression data from the Allen Human Brain Atlas (AHBA) (Figure 6). Across seven of the eight brain regions examined, i.e., the AMY, CN, CRB, HIP, NAc, PFC, and PUT, differentially methylated transcripts showed no significant enrichment in region-preferentially expressed genes (FDR > 0.05). In the VTA, a statistically significant result was observed (NES = −1.10, FDR = 2.87×10^-^³); however, the negative NES value indicates that differentially methylated transcripts were depleted from region-preferentially expressed genes rather than enriched, which is consistent with the overall pattern of no positive enrichment across regions. Taken together, these results demonstrate that AUD-associated m^6^A dysregulation is not confounded by regional gene expression signatures, but rather reflects disease-specific epitranscriptomic reprogramming in AUD.

**Fig. 6.**
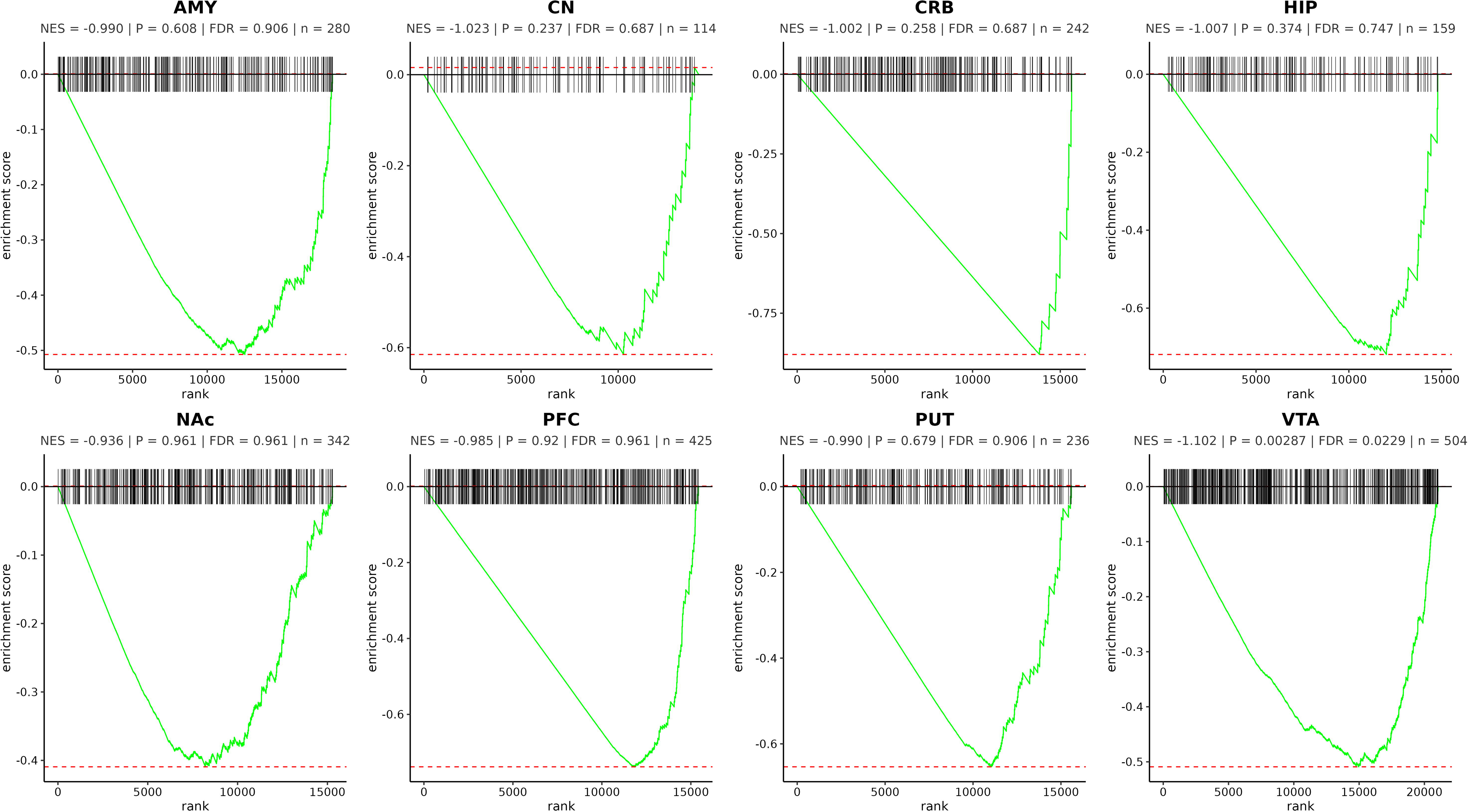
Enrichment analysis of differentially methylated transcripts in region-preferentially expressed genes. Each panel displays the running enrichment score (y-axis) across the ranked AHBA gene list (x-axis), with tick marks indicating the positions of differentially methylated transcripts within the ranked list. The normalized enrichment score (NES) and false discovery rate (FDR) are indicated within each panel. Statistically significant enrichments are defined as FDR < 0.05. Abbreviations: AMY, amygdala; CN, caudate nucleus; CRB, cerebellum; HIP, hippocampus; NAc, nucleus accumbens; prefrontal cortex; PUT, putamen; VTA, ventral tegmental area.

## DISCUSSION

This study presents the first systematic, multi-region epitranscriptomic survey of m^6^A RNA methylation in postmortem human brains from individuals with AUD, profiling 10,249 m^6^A sites across eight functionally distinct reward- and motor-related brain regions. Our principal findings are: (1) global m^6^A levels did not differ significantly between AUD cases and controls across any of the eight brain regions examined; (2) AUD was associated with site-selective differential m^6^A methylation at 1,403 unique sites distributed across all eight regions, with dysregulation predominantly region-specific in character; (3) differentially methylated transcripts converged on five functional categories, including cell adhesion and intercellular communication, growth-factor and morphogen signaling, immune and inflammatory signaling, RNA processing and protein homeostasis, and energy metabolism and neurodegeneration-associated pathways, despite marked regional heterogeneity in individual pathway membership; (4) the direction of m^6^A changes was consistently coupled with concordant gene expression alterations in a pattern consistent with canonical m^6^A-mediated mRNA decay; and (5) AUD-associated m^6^A dysregulation was not attributable to inherent regional gene expression identity, supporting a disease-specific epitranscriptomic reprogramming interpretation.

### Global versus site-specific m^6^A remodeling in AUD

The absence of statistically significant differences in global m^6^A levels between AUD cases and controls across all eight brain regions indicates that AUD-associated epitranscriptomic dysregulation operates through site-selective rather than global methylation remodeling. The hippocampus showed a trend toward elevated global m^6^A in AUD subjects (median = 0.049 vs. 0.039 in controls, *P* = 0.053) (Supplementary Figure 3), which is noteworthy given the established roles of the hippocampus in contextual memory encoding and cue-triggered craving, processes critically involved in relapses. Stress-induced elevation of global m^6^A in hippocampal tissues has been reported in rodent models ^42^, suggesting that this region may be particularly sensitive to experience-dependent epitranscriptomic changes. The trend observed here warrants follow-up in larger, adequately powered cohorts.

Our previous study reported increased global m^6^A levels following chronic intermittent ethanol (CIE) exposure in cultured cells ^32^. The discordance between cell-based global m^6^A elevation and the region-specific, site-selective pattern observed in human postmortem tissue likely reflects important differences in cellular context, exposure duration, and the heterogeneous composition of postmortem brain tissue, in which multiple neuronal and glial cell types with potentially opposing m^6^A responses are represented in bulk.

Collectively, these observations suggest that global m^6^A quantification lacks the sensitivity and resolution required to capture the full scope of alcohol-induced epitranscriptomic dysregulation in the human brain.

### Region-specific patterns of differential m^6^A methylation

Site-specific differential methylation was identified across all eight brain regions, with the PFC exhibiting the greatest number of dysregulated sites (n = 362) and the CN the fewest (n = 91) (Supplementary Figure 4). The prominence of PFC dysregulation is consistent with its central role in executive control, decision-making, and inhibitory regulation, i.e., the functions that are markedly compromised in AUD and have been associated with extensive transcriptional and DNA methylation alterations in prior studies ^5, 8, 19^. Our data extend these observations to the epitranscriptomic level, suggesting that post-transcriptional dysregulation through m^6^A may represent an additional and previously unappreciated layer of PFC molecular dysfunction in AUD. The substantial numbers of differentially methylated sites in the NAc (n = 276) and VTA (n = 241) are likewise consistent with the central importance of these mesolimbic regions in dopamine-mediated reinforcement, motivational salience, and reward-related plasticity. The VTA-NAc axis is particularly implicated in the transition from controlled to compulsive alcohol use, and epitranscriptomic dysregulation in this circuit may contribute to the maladaptive motivational states that perpetuate addictive behavior. The relatively lower site counts in the CN and HIP may reflect genuine region-specific differences in the extent of epitranscriptomic remodeling, differences in cellular composition affecting the sensitivity of bulk-tissue profiling, or residual variability attributable to the modest statistical power of the present study. Directional patterns were also region-dependent: the PFC and NAc exhibited an approximately equal balance of hyper- and hypomethylated sites, whereas the CRB was predominantly hypermethylated and the AMY predominantly hypomethylated, suggesting divergent regulatory responses across brain regions that may underlie their distinct contributions to AUD-associated behavioral and neurological sequelae.

### Predominance of region-specific m^6^A dysregulation

A defining feature of our results was the overwhelming predominance of region-specific m^6^A dysregulation: 86.5% of the 1,403 differentially methylated sites were unique to a single brain region, and no site was shared across more than three of the eight regions examined. This extensive regional specificity is consistent with the distinct functional architecture, cellular composition, and transcriptional landscape of these brain regions and indicates that AUD-associated epitranscriptomic remodeling is not a uniform, brain-wide phenomenon but rather a spatially patterned molecular response. Region-specific epigenetic heterogeneity in AUD has been documented at the DNA methylation level across multiple postmortem cohorts ^5–7^, and our transcriptomic analyses similarly revealed region-specific differential expression patterns across the same eight brain regions ^19^. The convergence of region-specific dysregulation across DNA methylation, transcriptomic, and now epitranscriptomic datasets supports the view that AUD induces heterogeneous, region-tailored molecular adaptations that may collectively underlie the heterogeneous clinical manifestations of the disorder, including the distinct profiles of cognitive impairment, affective dysregulation, and motor dysfunction that characterize different stages and severities of AUD.

### Biological significance of pathway enrichment

Pathway enrichment analysis revealed that differentially methylated transcripts converged on five broad functional systems, despite heterogeneity in the specific enriched pathways across individual brain regions. The enrichment of cell adhesion and intercellular communication pathways, including *Adherens Junction*, *Gap Junction*, *Focal Adhesion*, and *ECM-receptor Interactio*n, across multiple regions suggests that m^6^A-mediated post-transcriptional dysregulation of synaptic structure and glial-neuronal communication may contribute to the synaptic remodeling that characterizes chronic alcohol exposure. Cell adhesion molecules are critical determinants of synapse formation, stabilization, and plasticity ^43^, and their disruption at the epitranscriptomic level may complement the well-documented transcriptional dysregulation of synaptic genes in AUD ^8, 19^.

The enrichment of growth-factor and morphogen signaling pathways, including *ErbB*, *RTK-RAS-ERK*, *RTK-RAS-PI3K*, *TGF-*β, and *Wnt Signaling*, is consistent with established roles of these cascades in neuronal survival, dendritic arborization, synaptic plasticity, and neurogenesis, all of which are compromised in AUD. These pathways have been identified as transcriptional targets of alcohol-induced epigenetic dysregulation in prior studies ^8, 11^, and the present data indicate that post-transcriptional m^6^A-mediated regulation of these pathways provides an additional, potentially faster-acting layer of alcohol-induced molecular control that operates in parallel with transcriptional mechanisms.

The enrichment of immune and inflammatory pathways, including *RIG-I-like Receptor Signaling*, *Antigen Processing and Presentation*, *NK Cell-mediated Cytotoxicity*, and *Adipocytokine Signaling*, is consistent with the well-established role of neuroinflammation in AUD pathophysiology. Innate immune activation in the brain, driven in part by alcohol-induced Toll-like receptor signaling and microglial activation, has been extensively implicated in the neuropathology, mood dysregulation, and cognitive deficits associated with AUD ^44^. m^6^A has emerged as a key regulator of innate immune transcript stability and translational efficiency, modulating the post-transcriptional fate of cytokine and interferon-stimulated gene mRNAs ^45^, and our data suggests that m^6^A-mediated dysregulation of immune transcripts may contribute to the sustained neuroinflammatory state in AUD brains.

The enrichment of RNA processing and protein homeostasis pathways, including *Ribosome*, *Translation Initiation*, *Spliceosome*, and *Ubiquitin-mediated Proteolysis*, indicates that AUD-associated m^6^A dysregulation extends to the machinery governing gene expression itself. Disruption of translational capacity and protein quality control systems may contribute to the synaptic dysfunction and progressive neuronal vulnerability observed in severe AUD. The enrichment of *Histone H2AK119 Deubiquitination* in the hippocampus further links m^6^A dysregulation to chromatin-level regulatory mechanisms, suggesting crosstalk between the epitranscriptome and the epigenome in AUD.

Finally, the enrichment of energy metabolism and neurodegeneration-associated pathways, including *ADRB3-UCP1 Signaling*, *Vasopressin-regulated Water Reabsorption*, and the *Parkinson’s* and *Huntington’s Disease* pathway gene sets, is consistent with the metabolic dysregulation and progressive neurotoxicity associated with chronic alcohol exposure ^46^. The overlap with neurodegeneration-associated pathways supports the notion that the molecular sequelae of AUD shares mechanistic features with neurodegenerative processes, possibly reflecting convergent dysregulation of mitochondrial function, proteostasis, and neuroinflammatory signaling.

### m^6^A dysregulation is mechanistically coupled with altered gene expression

A central and mechanistically informative finding of this study is the directional coupling between site-specific m^6^A changes and transcript abundance: hypermethylated transcripts were preferentially enriched among downregulated genes in the AUD transcriptome (significant in AMY, CN, and PUT), while hypomethylated transcripts were enriched among upregulated genes (significant in CN, CRB, and PUT) (Figures 4 and 5). This consistent bidirectional pattern is concordant with the canonical role of m^6^A in promoting mRNA degradation through recognition by YTH domain-containing reader proteins, particularly YTHDF2, which recruits the CCR4-NOT deadenylase complex to facilitate poly(A) tail shortening and mRNA decay ^47, 48^. The mechanistic coherence of these findings provides strong support for m^6^A remodeling as a post-transcriptional regulatory layer that directly contributes to, and likely reinforces, the transcriptional dysregulation previously documented in AUD brains. The regional heterogeneity in the statistical significance of this coupling, i.e., significant in some regions but not others, may reflect differences in the expression levels or activity of YTHDF reader proteins across brain regions, variation in the proportion of m^6^A-bearing transcripts subject to degradation-promoting versus translation-enhancing reader recognition, or simply insufficient statistical power in regions with fewer differentially methylated transcripts.

### AUD-specific epitranscriptomic reprogramming: Evidence from Allen Human Brain Atlas integration

The AHBA integrative analysis provided critical evidence that the observed m^6^A dysregulation reflects disease-specific changes rather than confounding by inherent regional gene expression signatures. In seven of the eight brain regions, differentially methylated transcripts showed no significant enrichment among region-preferentially expressed genes, indicating that AUD-associated m^6^A alterations are not simply recapitulating the transcriptional identity of each brain region. In the VTA, the statistically significant result indicated depletion of differentially methylated transcripts from region-preferentially expressed genes rather than enrichment, suggesting that AUD-associated m^6^A changes in the VTA preferentially affect transcripts not characteristic of VTA cellular identity. The VTA is a small, neurochemically heterogeneous region comprising dopaminergic, GABAergic, and glutamatergic neurons, and the observed pattern may reflect selective epitranscriptomic dysregulation of transcripts specific to VTA cell subtypes, an interpretation that future single-nucleus analyses will be well positioned to address. Collectively, these analyses strengthen the conclusion that the m^6^A alterations reported here represent genuine AUD-driven epitranscriptomic reprogramming rather than a secondary consequence of regional transcriptional architecture.

### Limitations

Several important limitations must be acknowledged when interpreting the present findings. First, the cross-sectional postmortem design inherently precludes causal inference: it is not possible to determine whether the identified m^6^A alterations precede AUD development as a predisposing factor, emerge as molecular consequences of chronic alcohol exposure, or reflect a combination of both. Second, the relatively low mean RIN (4.8 ± 1.4) is a recognized constraint of archived postmortem tissue; however, the MazF-based assay was selected specifically for its tolerance of RNA fragmentation, and spike-in normalization was implemented to minimize RNA quality-related technical noise. Third, while the sample size (n = 12 per group) is consistent with postmortem brain studies of this nature, statistical power was limited, and the use of nominal significance thresholds (*P* < 0.05, |FC| ≥ 1.5) without FDR correction at the site level reflects this constraint; accordingly, many of the reported sites should be considered candidates requiring replication in larger, independently ascertained cohorts. Fourth, the cohort comprised exclusively Caucasian Australians, limiting the generalizability of findings to populations of different genetic ancestries. Fifth, residual confounding from comorbid psychiatric diagnoses, polydrug use, smoking, and liver disease cannot be fully excluded despite attempts to account for these variables during cohort curation. Sixth, because bulk tissue was profiled, cell-type-specific m^6^A dynamics are unresolved; alterations detected in bulk may represent aggregate signals from multiple neuronal and glial populations, potentially masking cell-type-specific effects of greater biological magnitude. Finally, the Arraystar m^6^A Single Nucleotide Array covers validated m^6^A sites at ACA motifs and does not provide transcriptome-wide coverage; m^6^A changes at sites outside the array’s probe set, particularly at non-ACA motifs, remain uncharacterized.

### Conclusions

In summary, this study provides the first comprehensive, multi-region characterization of m^6^A RNA epitranscriptomic dysregulation in human AUD postmortem brains. We demonstrate that AUD is associated with predominantly region-specific, site-selective m^6^A remodeling that is likely coupled to altered transcript abundance through the canonical m^6^A-mediated mRNA decay axis. The convergence of dysregulated transcripts on intercellular communication, neuroinflammatory signaling, RNA and protein processing, growth factor cascades, and metabolic pathways, despite extensive regional heterogeneity at the level of individual sites, reveals that m^6^A remodeling constitutes a pervasive and functionally consequential layer of post-transcriptional regulation in AUD. The specificity of these changes for AUD pathology, as demonstrated by the AHBA integrative analysis, underscores their biological and translational relevance. Collectively, these findings position the m^6^A epitranscriptome as a novel and underappreciated dimension of molecular plasticity in AUD, providing a foundation for future investigations into epitranscriptomic biomarkers and the potential therapeutic targeting of m^6^A regulatory machinery in AUD.

## ACKNOWLEDGMENTS

This work was supported by the National Institute on Alcohol Abuse and Alcoholism (NIAAA), R01 AA029758. We thank the New South Wales Brain Tissue Resource Centre (NSWBTRC) at the University of Sydney for providing AUD and control postmortem brain tissue; the NSWBTRC is supported by the University of Sydney, the National Health and Medical Research Council of Australia, and the NIAAA. We are grateful to the donors and their next of kin for tissue donation and consent for research use. We also thank the Arraystar team for performing the m^6^A microarray experiments.

## AUTHOR CONTRIBUTIONS

HZ conceived and designed the study. HZ, AK, and OTOB contributed substantially to manuscript writing and drafting. HZ and JSK performed tissue dissection and sample preparation. HZ, AK, OTOB, ASP, AP, YK, and JEB conducted data analysis and contributed to interpretation of results. HZ secured funding for the project. All authors contributed to manuscript revision, approved the final version, and agreed to be accountable for all aspects of the work.

## ETHICS APPROVAL

This study was conducted in accordance with institutional biosafety and ethical standards and was approved by the Boston University Institutional Biosafety Committee (Project Approval No. 23 2264). Use of human postmortem tissue was performed under the New South Wales Brain Tissue Resource Centre’s governance and donor consent procedures; any applicable data use agreements and donor protections were observed.

## DATA AVAILABILITY

The microarray data supporting the findings of this study are available from the corresponding author upon reasonable request.

## ADDITIONAL INFORMATION

Supplementary information is available at MP’s website.

