## Supplemental Materials for "Altered m^6^A methylation across eight reward- and motor-related brain regions in alcohol use disorder"

**(Supplementary Information)**

**Supplementary Fig. 1.** Global m^6^A RNA methylation quantification.

**Supplementary Fig. 2.** PCA before and after QC and batch effect correction.

**Supplementary Fig. 3.** Global m^6^A methylation differences across eight brain regions between AUD and control subjects.

**Supplementary Fig. 4.** The counts of hyper- and hypomethylated sites in each brain region.

**Supplementary Fig. 5.** The counts of transcripts with exclusively hyper-, exclusively hypo-, or both hyper- and hypomethylated sites in each brain region.

**Supplementary Fig. 6.** Chromosome circos diagrams illustrating the distributions of differentially methylated m^6^A sites across the eight brain regions.

**Supplementary Table 1** Demographic summary of postmortem brain samples by diagnosis

**Supplementary Table 2** Top 10 differentially methylated m^6^A sites (with smallest *P* values) in each brain region of AUD subjects

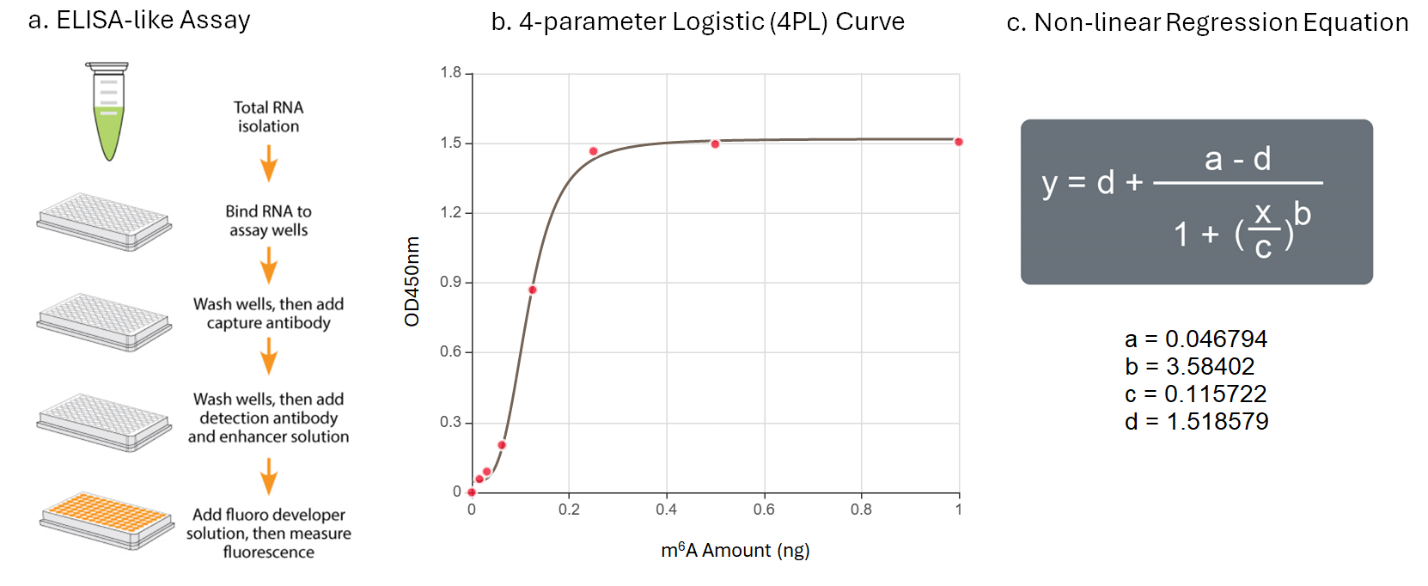

**Supplementary Fig. 1.** Global m^6^A RNA methylation quantification.

a. ELISA-like assays; b: 4-parametric logistic (4PL) curve; c: Non-linear regression equation

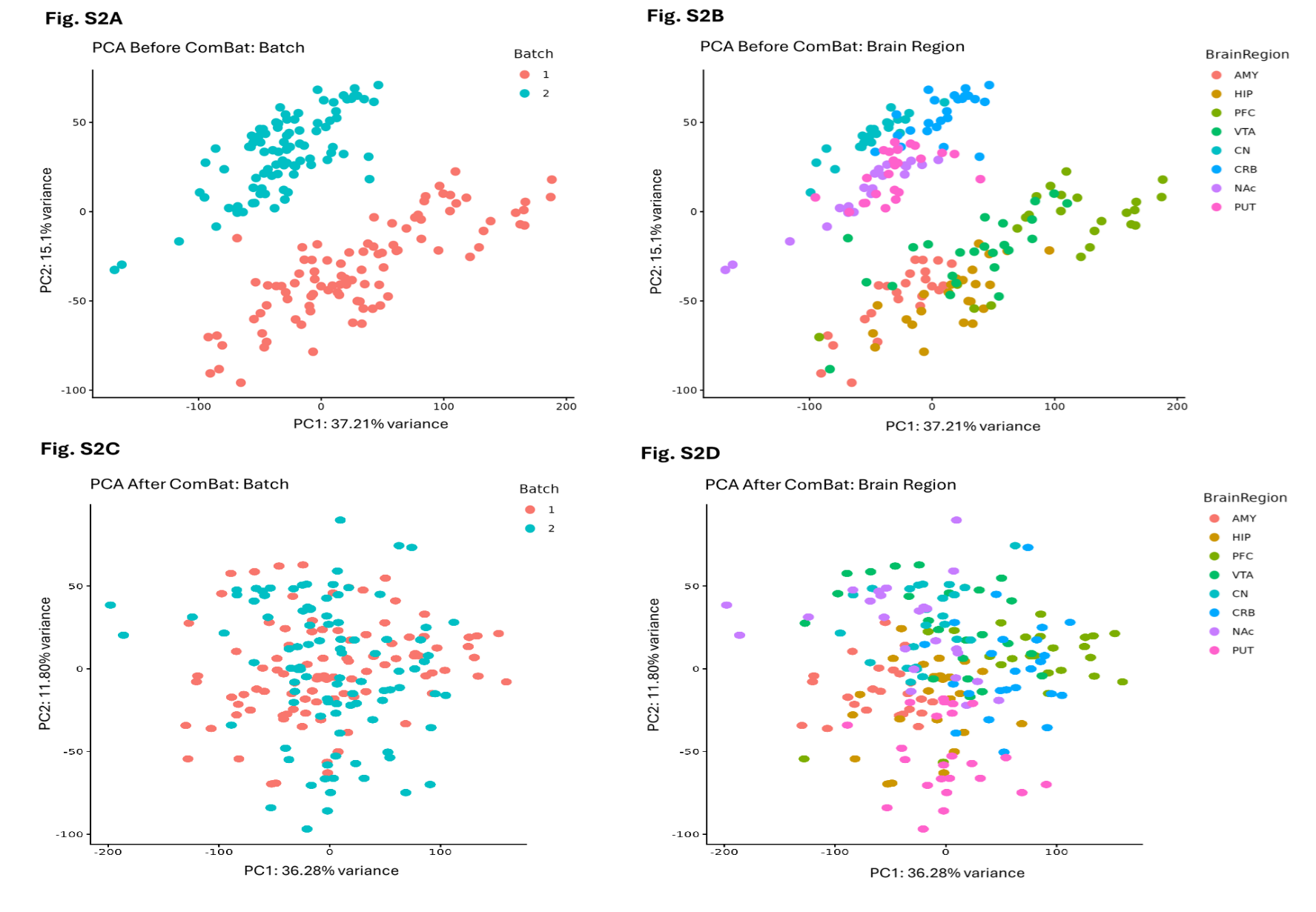

**Supplementary Fig. 2.** PCA before and after QC and batch effect correction.

Fig. S2A and Fig. S2C: PCA by batches (Batch 1: 96 samples; Batch 2: 96 samples); Fig. S2B and Fig. S2D: PCA by brain regions [Batch 1: AMY (amygdala), HIP (hippocampus), PFC (prefrontal cortex), and VTA (ventral tegmental area); Batch 2: CN (caudate nucleus), CRB (cerebellum), NAc (nucleus accumbens), and PUT (putamen)].

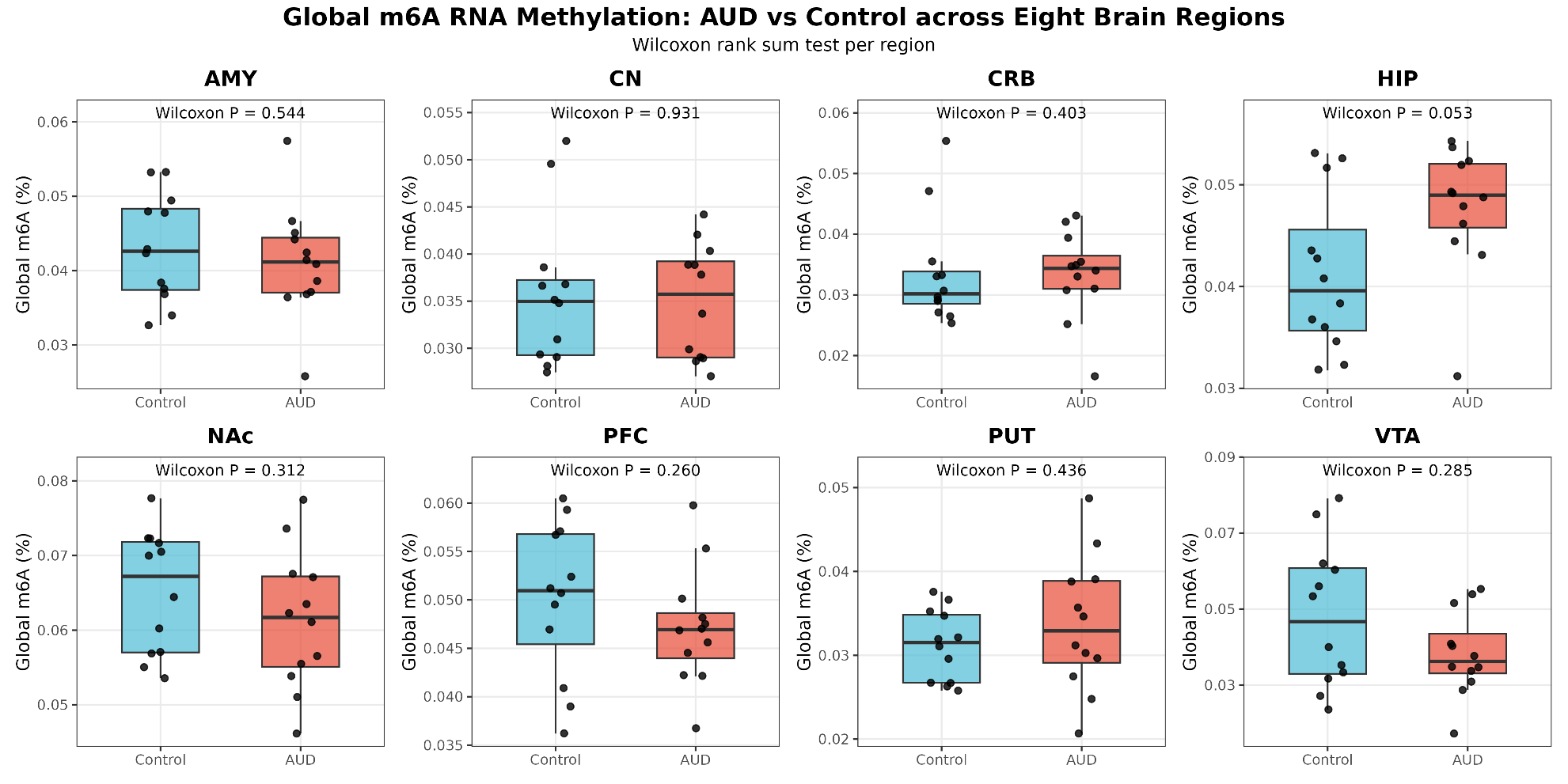

**Supplementary Fig. 3** Global m^6^A methylation differences across eight brain regions between AUD and control subjects.

AMY (amygdala), CN (caudate nucleus), CRB (cerebellum), HIP (hippocampus), NAc (nucleus accumbens), PFC (prefrontal cortex), PUT (putamen), and VTA (ventral tegmental area).

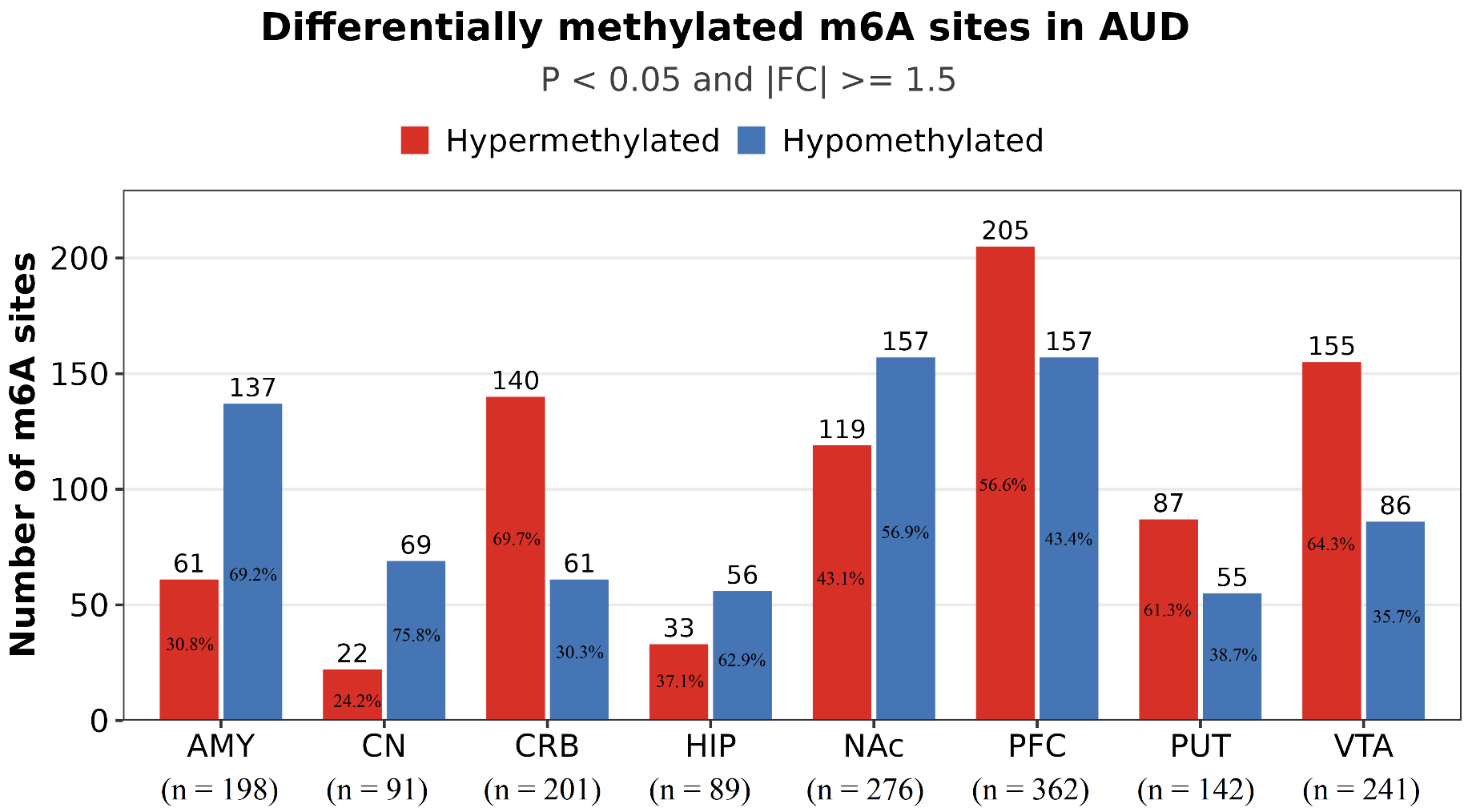

**Supplementary Fig. 4.** The counts of hyper- and hypomethylated sites in each brain region.

Bar chart showing the number of differentially methylated m^6^A sites identified in each of the eight brain regions in individuals with alcohol use disorder (AUD) compared to matched controls. Hypermethylated sites (shown in red) and hypomethylated sites (shown in blue) are displayed as separate bars for each region. Differential methylation was defined by an unadjusted *P* < 0.05 and an absolute fold change ≥ 1.5. Brain regions analyzed include the amygdala (AMY), caudate nucleus (CN), cerebellum (CRB), hippocampus (HIP), nucleus accumbens (NAc), prefrontal cortex (PFC), putamen (PUT), and ventral tegmental area (VTA).

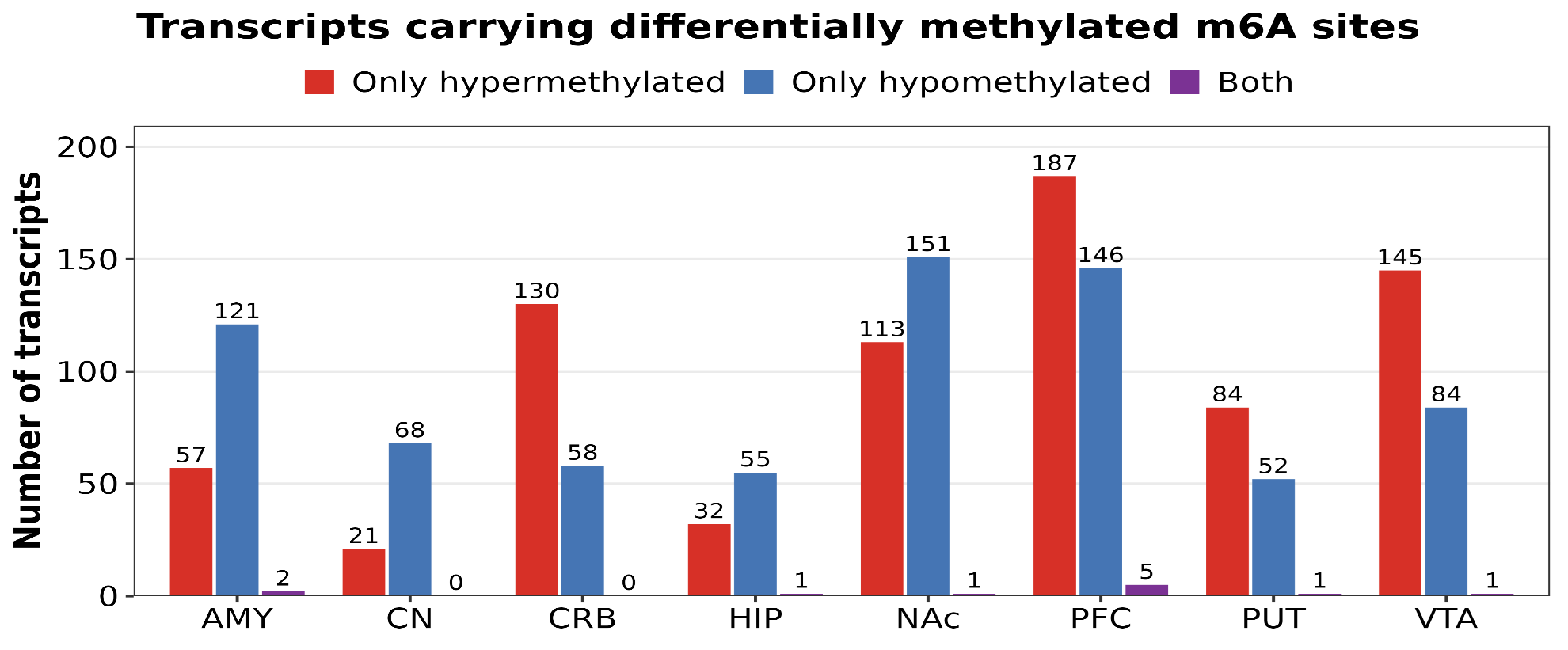
**Supplementary Fig. 5.** The counts of transcripts with exclusively hyper-, exclusively hypo-, or both hyper- and hypomethylated sites in each brain region.

Grouped bar chart showing the number of transcripts categorized by their m^6^A methylation pattern in each of the eight brain regions in individuals with alcohol use disorder (AUD) compared to matched controls. For each brain region, transcripts are classified into three categories: exclusively hypermethylated (containing only hypermethylated m^6^A sites; red), exclusively hypomethylated (containing only hypomethylated m^6^A sites; blue), and both hyper- and hypomethylated (containing at least one hypermethylated and at least one hypomethylated m^6^A site; purple). Differentially methylated sites were identified using an unadjusted *P* < 0.05 and an absolute fold change ≥ 1.5. Brain regions analyzed include the amygdala (AMY), caudate nucleus (CN), cerebellum (CRB), hippocampus (HIP), nucleus accumbens (NAc), prefrontal cortex (PFC), putamen (PUT), and ventral tegmental area (VTA).

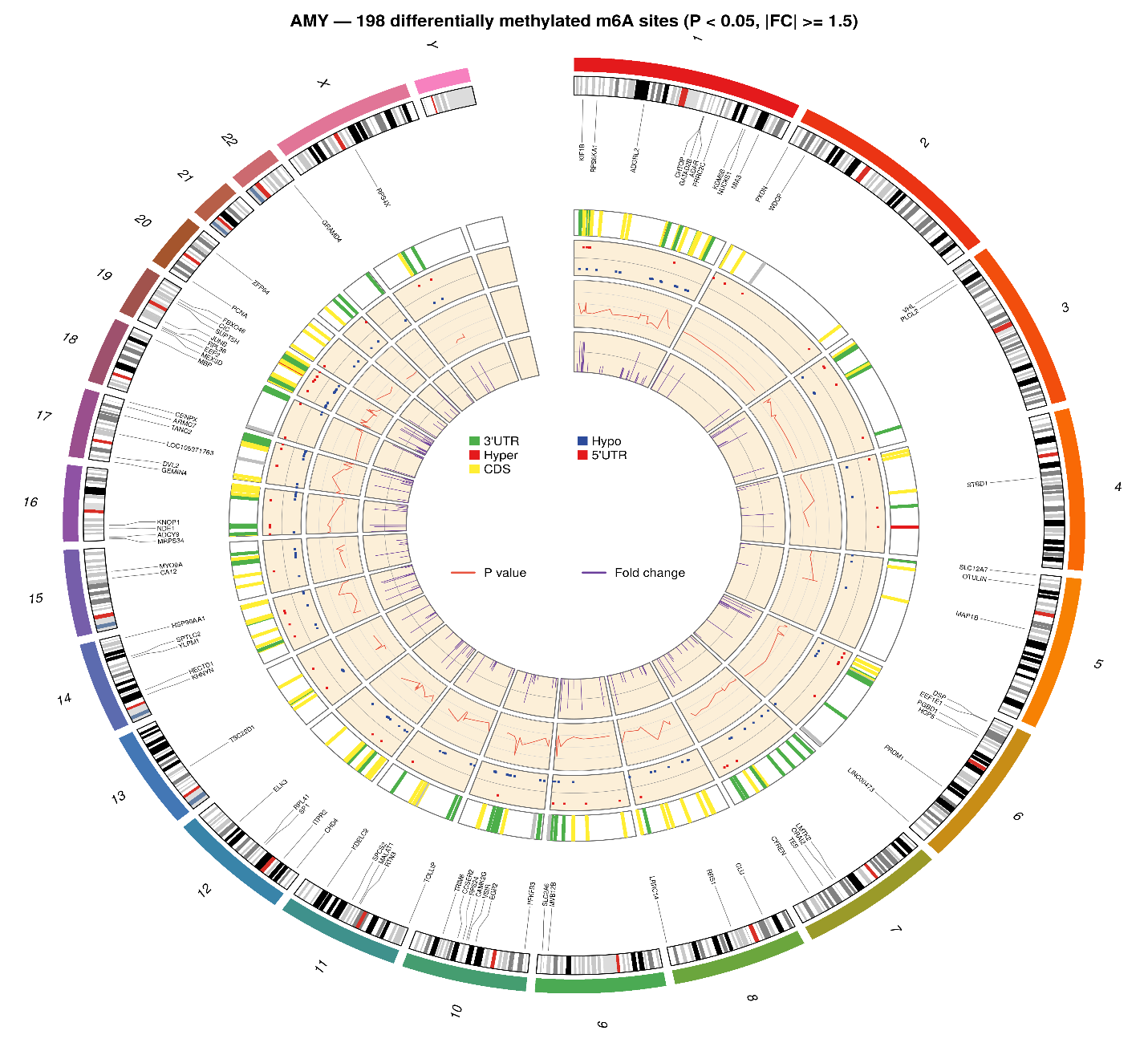

**Supplementary Fig. S6a (Amygdala)**

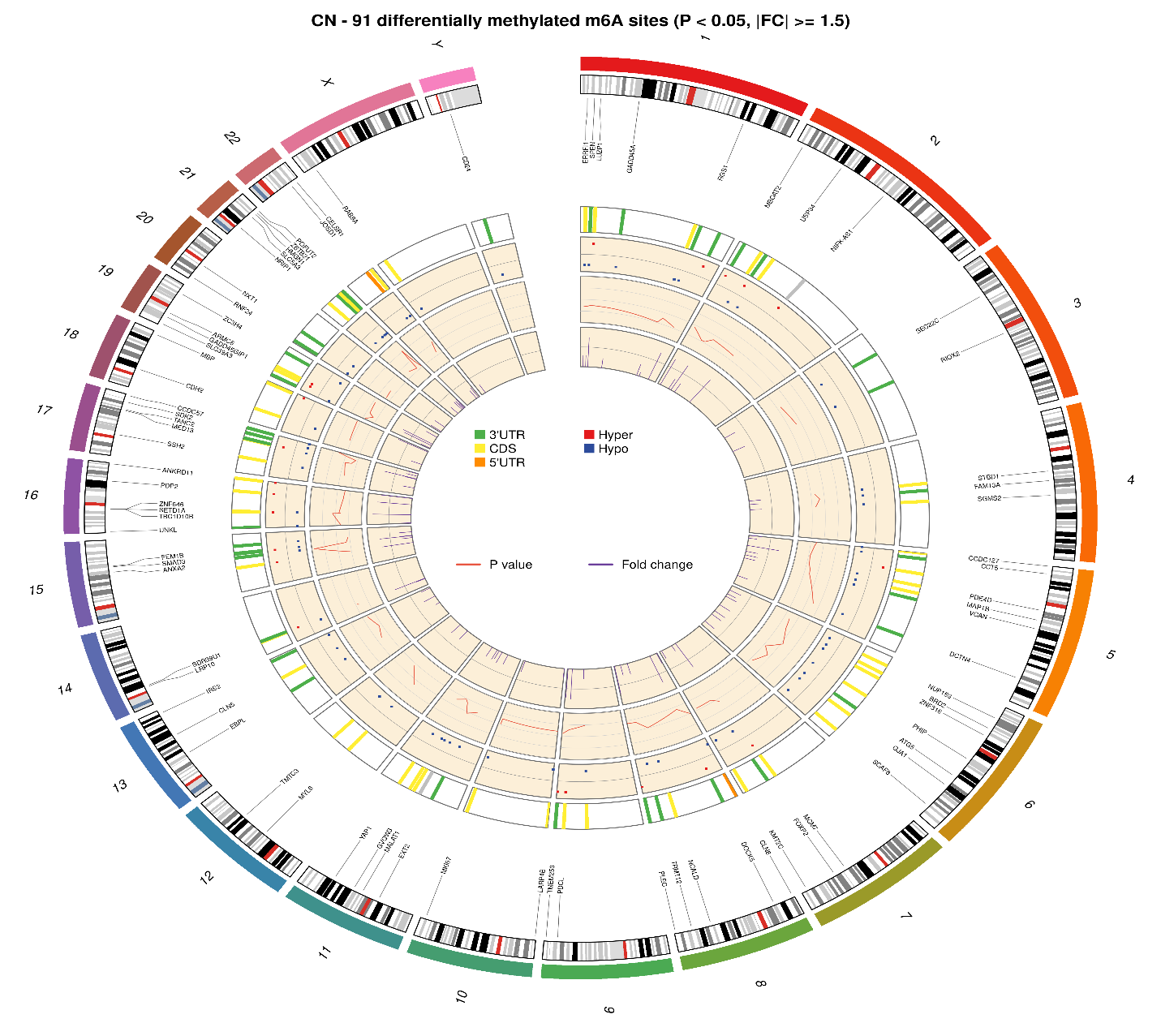

**Supplementary Fig. S6b (Caudate Nucleus)**

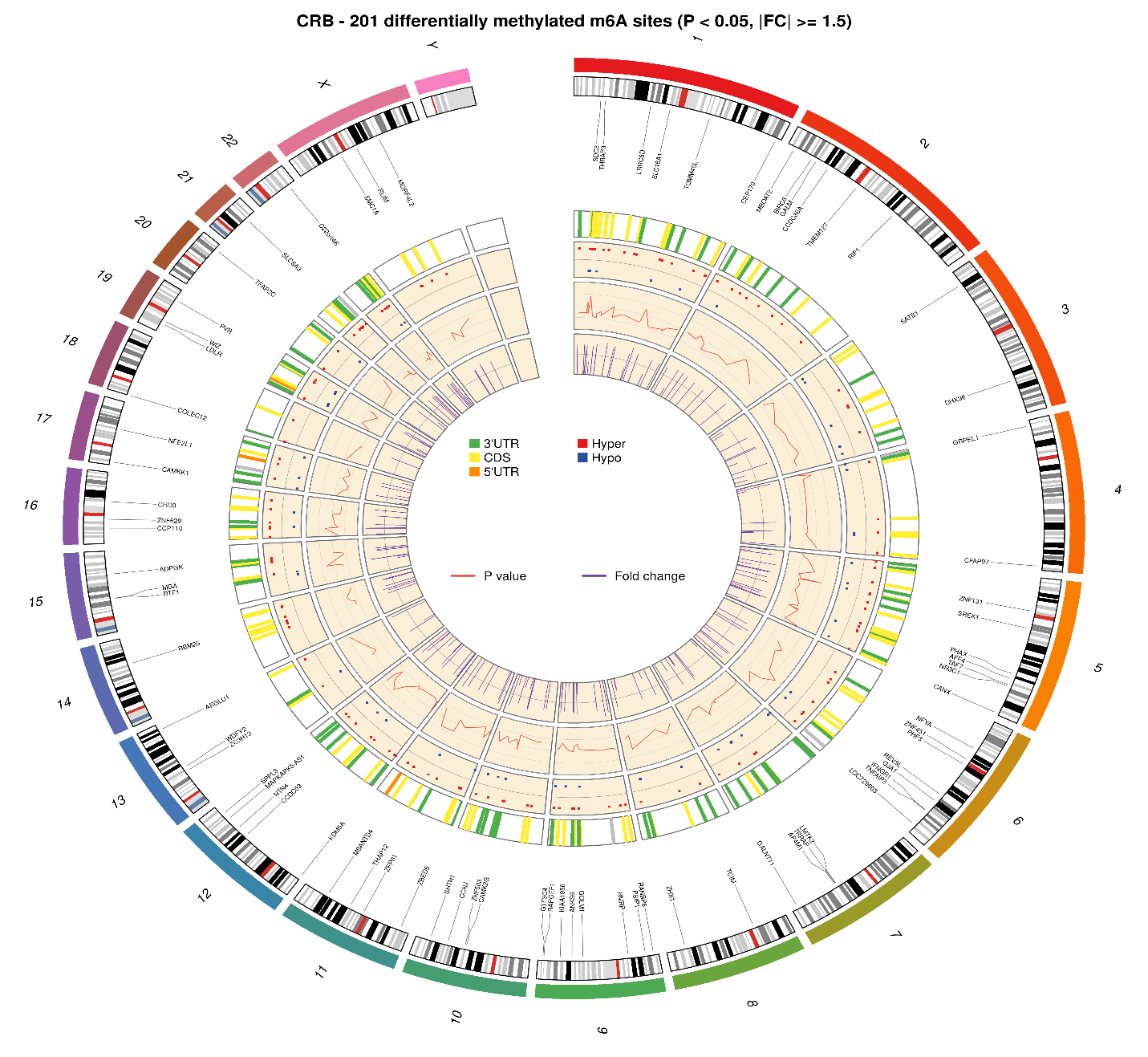

**Supplementary Fig. S6c (Cerebellum)**

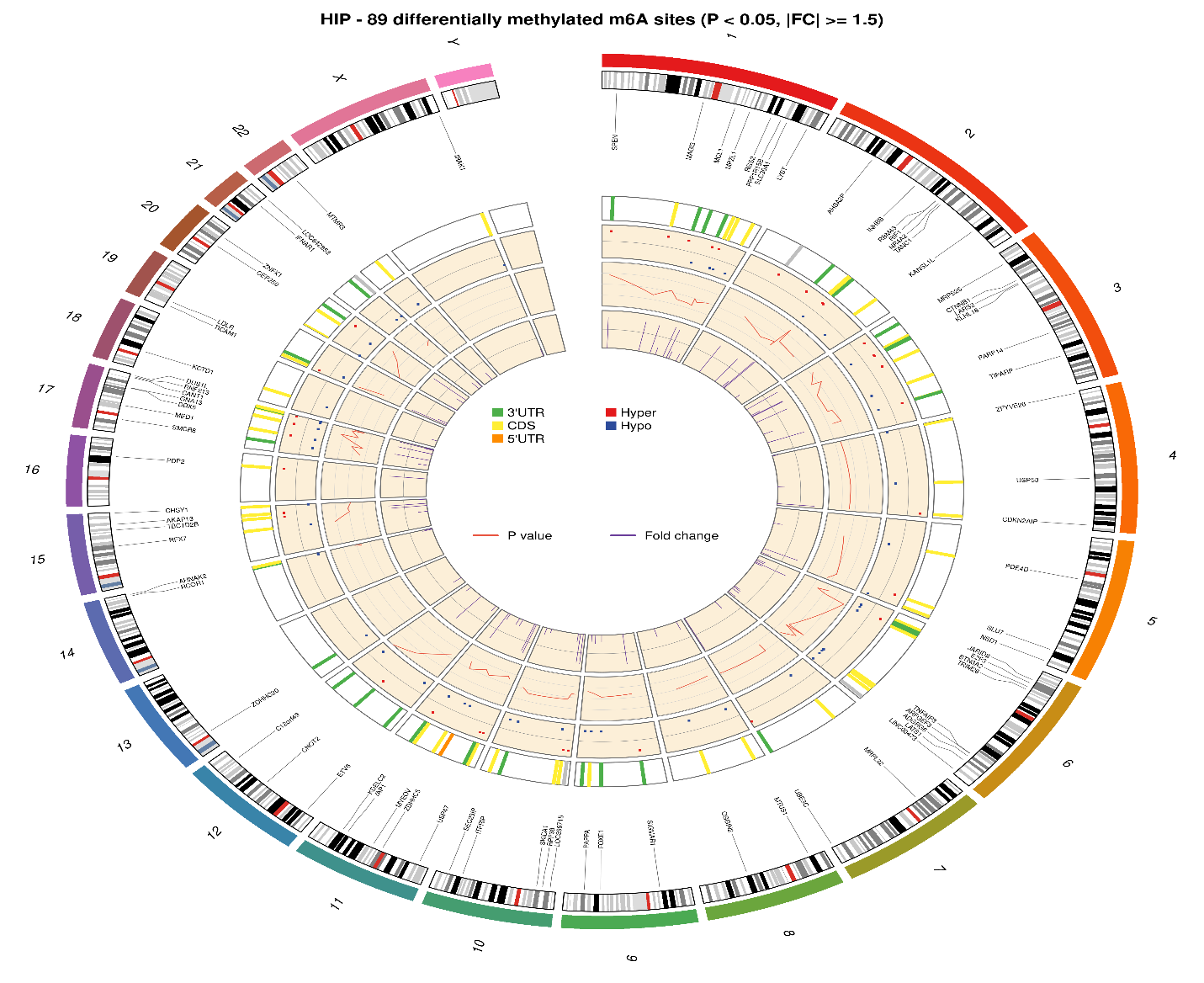

**Supplementary Fig. S6d (Hippocampus)**

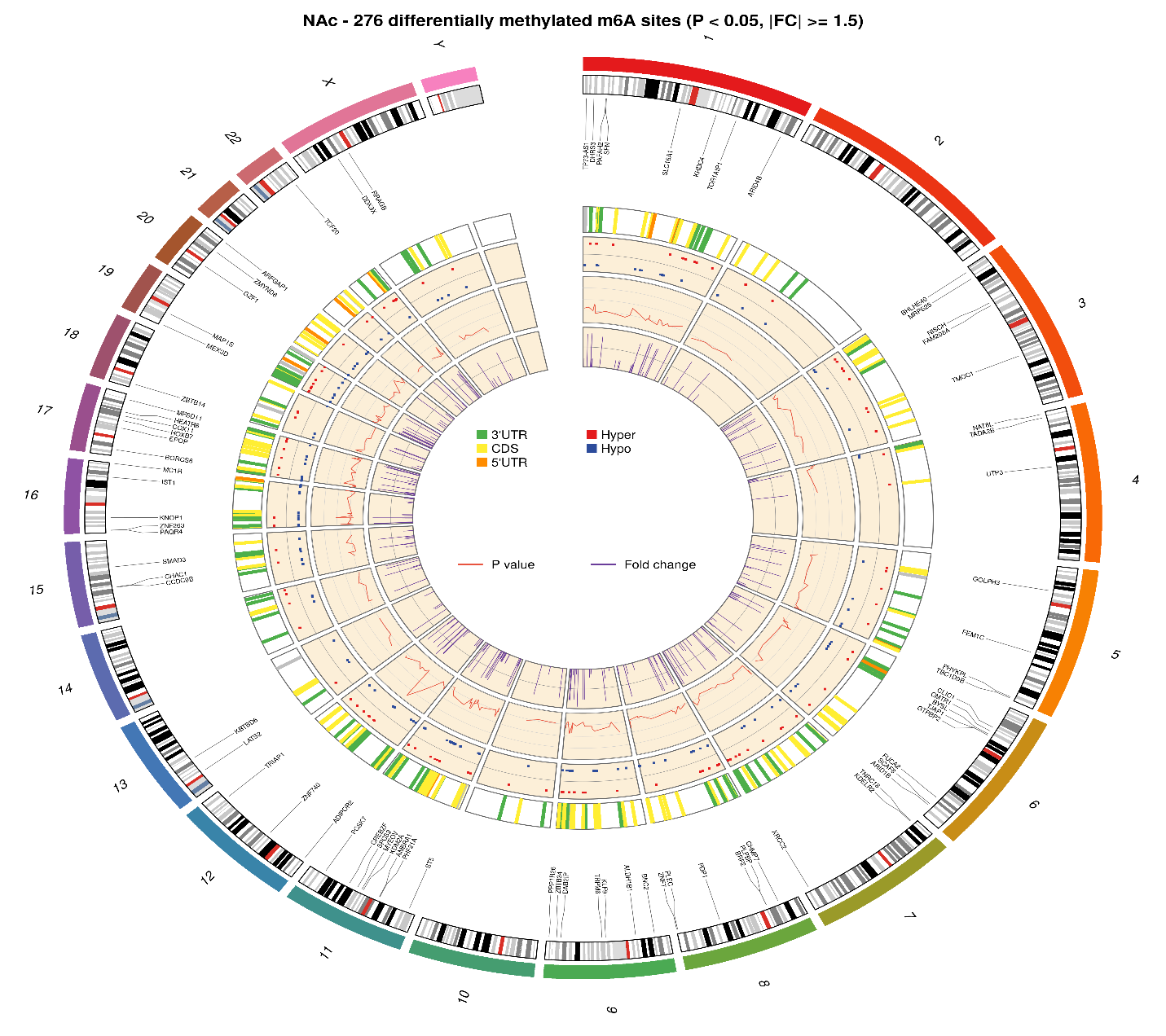

**Supplementary Fig. S6e (Nucleus Accumbens)**

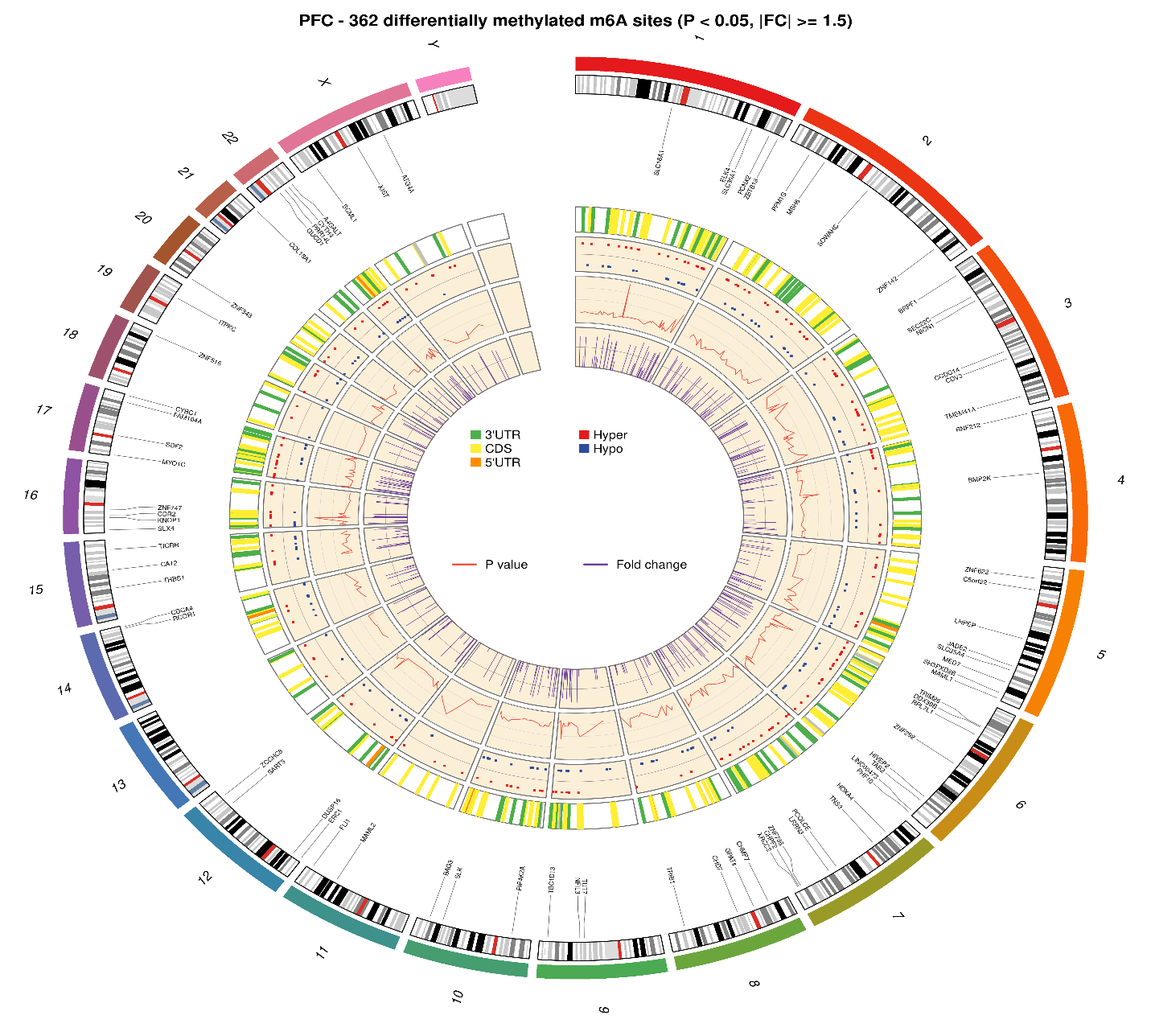

**Supplementary Fig. S6f (Prefrontal Cortex)**

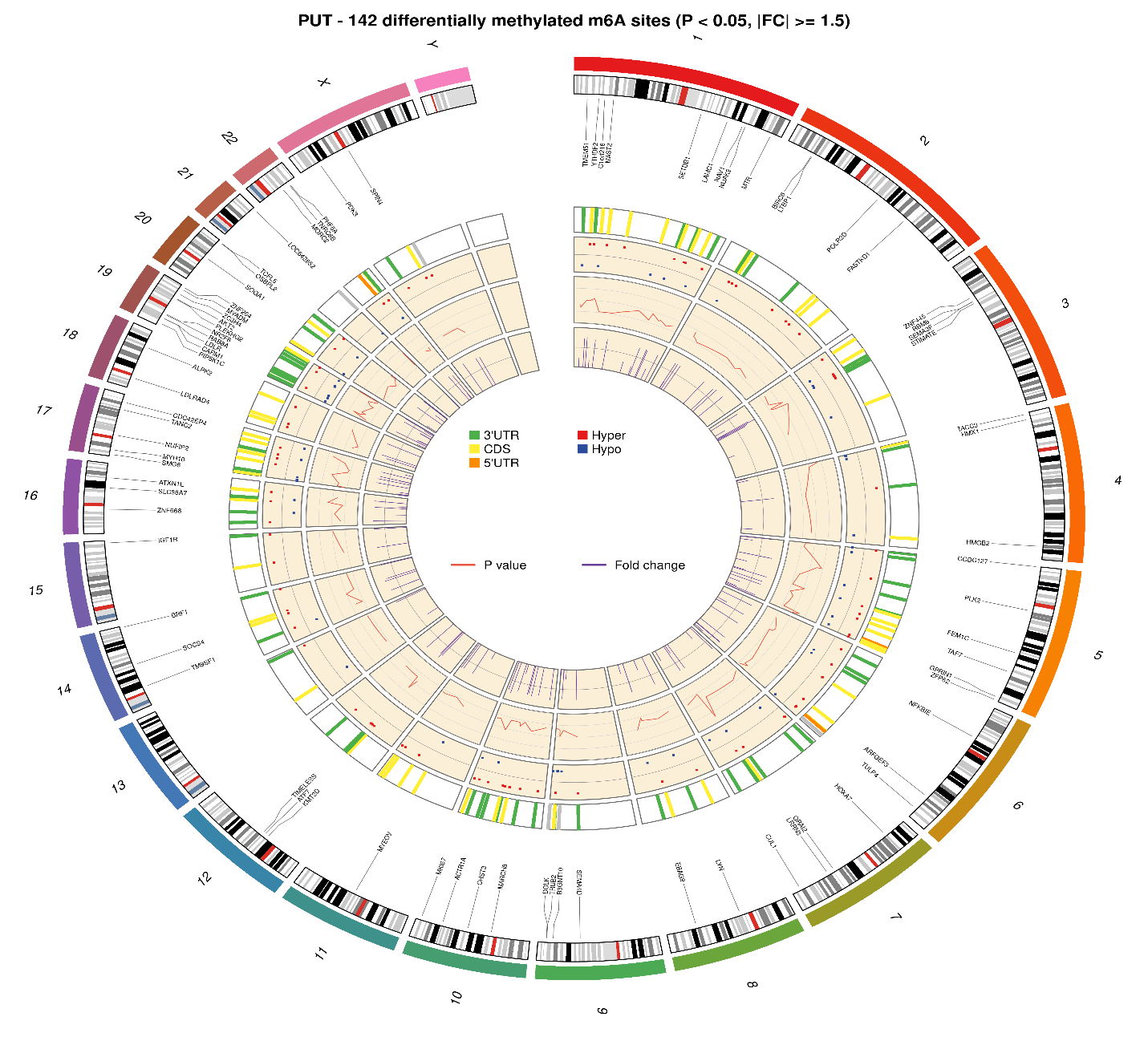

**Supplementary Fig. S6g (Putamen)**

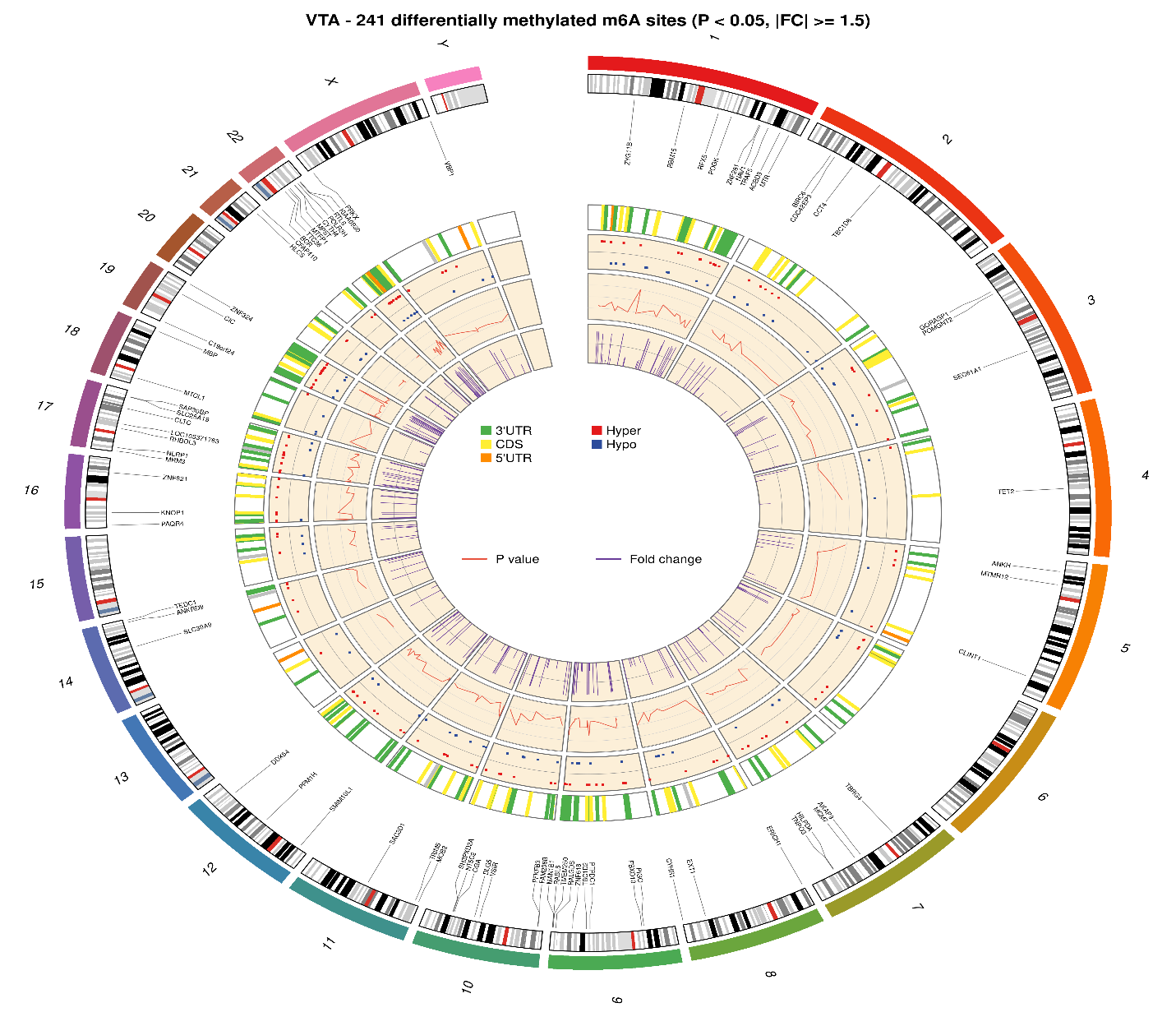

**Supplementary Fig. S6h (Ventral Tegmental Area)**

**Fig. S6.** Chromosome circos diagrams illustrating the distributions of differentially methylated m^6^A sites across the eight brain regions.

Circos diagrams depicting the chromosomal distribution of differentially methylated m^6^A sites (*P* < 0.05 & |FC| ≥ 1.5) identified in each of the eight brain regions in individuals with alcohol use disorder (AUD) compared to matched controls. Each panel represents one brain region: amygdala (AMY; Fig. S6a), caudate nucleus (CN: Fig. S6b), cerebellum (CRB: Fig. S6c), hippocampus (HIP: Fig. S6d), nucleus accumbens (NAc: Fig. S6e), prefrontal cortex (PFC; Fig. S6f), putamen (PUT: Fig. S6g), and ventral tegmental area (VTA: Fig. S6h). Within each diagram, rings are ordered from outermost to innermost as follows: chromosomal location, transcript names and positions, hyper- or hypomethylated sites, *P* values, and fold changes.

**Supplementary Table 1** Demographic summary of postmortem brain samples by diagnosis

| Clinical Data | AUD Cases | Controls | *P-*value |
| --- | --- | --- | --- |
| Daily alcohol use in grams (Mean±SD) | 336.9 (±405.6) | 9.6 (±10.4) | 0.010 |
| Sex | 6 (M), 6 (F) | 6 (M), 6 (F) | > 0.05 |
| Age (Mean±SD) | 48.4 (±6.6) | 50.2 (±11.2) | 0.645 |
| Postmortem interval in hours (Mean±SD) | 39.5 (±14.0) | 25.1 (±10.4) | 0.009 |
| RNA integrity number (Mean±SD) | AMY: 3.4 (±1.0) | AMY: 3.8 (±1.5) | 0.435 |
|  | CN: 5.1 (±1.4) | CN: 4.7 (±1.2) | 0.486 |
|  | CRB: 6.3 (±0.9) | CRB: 6.3 (±0.8) | 0.813 |
|  | HIP: 5.0 (±1.0) | HIP: 5.1 (±0.9) | 0.811 |
|  | NAc: 5.4 (±1.2) | NAc: 4.7 (±0.9) | 0.137 |
|  | PFC: 5.1 (±1.5) | PFC: 5.0 (±1.6) | 0.974 |
|  | PUT: 5.3 (±0.9) | PUT: 5.3 (±0.8) | 1.000 |
|  | VTA: 3.4 (±1.1) | VTA: 3.6 (±1.1) | 0.762 |
| Brain weight in grams (mean±SD) | 1407.3 (±190.3) | 1416.7 (±165.6) | 0.422 |
| Brain pH (mean±SD) | 6.6 (±0.3) | 6.6 (±0.3) | 0.924 |
| Cerebral hemispheres | Left: 5; Right: 7 | Left: 9; Right: 3 | > 0.05 |
| Smoking | Current: 9 | Current: 5 | > 0.05 |
|  | Never or former: 3 | Never or former: 7 |  |
| Liver disease | Yes: 9 | Yes: 5 | >0.05 |
|  | No: 3 | No: 7 |  |

AUD: alcohol use disorder.

AMY: Amygdala; CN: Caudate Nucleus; CRB: Cerebellum; HIPPO: Hippocampus; NAc: Nucleus Accumbens; PFC: Prefrontal Cortex; PUT: Putamen; and VTA: Ventral Tegmental Area.

Liver disease: with (Yes) or without (No) Steatosis or Congestion.

*P* values were calculated by t-tests for continuous variables and Fisher’s exact tests for categorical variables.

**Supplementary Table 2** Differentially methylated m^6^A sites (*P* < 0.05 & |logFC| ≥ 1.5; 10 sites with smallest *P* values) in each brain region of AUD subjects

| Brain_Region/  Gene_Symbol | mRNA/  ncRNA | Probe  Name | m^6^A  Location | AveMeth | Log_2_FC | t | B | *P*-value | *P*_adj_ |
| --- | --- | --- | --- | --- | --- | --- | --- | --- | --- |
| **Amygdala (AMY)** |  |  |  |  |  |  |  |  |  |
| *CAMK2G* | protein_coding | CAMK2G_1002520224 | 3'UTR | -3.23 | -1.06 | -3.99 | -3.12 | 9.7×10^-5^ | 0.821 |
| *NUCKS1* | protein_coding | NUCKS1_1003474544 | CDS | -6.32 | -0.88 | -3.75 | -3.29 | 0.0002 | 0.821 |
| *SLC2A6* | protein_coding | SLC2A6_1006452424 | 3'UTR | -3.01 | 0.85 | 3.72 | -3.31 | 0.0003 | 0.821 |
| *MAP1B* | protein_coding | MAP1B_1003900744 | CDS | -8.00 | -0.79 | -3.55 | -3.43 | 0.0005 | 0.821 |
| *RRS1* | protein_coding | RRS1_1001347784 | CDS | -2.72 | -0.78 | -3.42 | -3.51 | 0.0008 | 0.821 |
| *LOC105371763* | ncRNA | LOC105371763_1000329214 | 985 | -2.25 | 0.84 | 3.35 | -3.56 | 0.0010 | 0.821 |
| *SPTLC2* | protein_coding | SPTLC2_1000779054 | CDS | -3.25 | 0.69 | 3.34 | -3.57 | 0.0010 | 0.821 |
| *SPTLC2* | protein_coding | SPTLC2_1000652954 | 3'UTR | -3.95 | 0.78 | 3.34 | -3.57 | 0.0010 | 0.821 |
| *MAP1B* | protein_coding | MAP1B_1002914334 | CDS | -7.16 | -0.85 | -3.30 | -3.59 | 0.0011 | 0.821 |
| *MAP1B* | protein_coding | MAP1B_1001859354 | CDS | -7.05 | -0.88 | -3.29 | -3.60 | 0.0012 | 0.821 |
| **Caudate Nucleus (CN)** |  |  |  |  |  |  |  |  |  |
| *ANXA2* | protein_coding | ANXA2_1002215614 | 3'UTR | -3.79 | -0.73 | -4.41 | -2.83 | 1.8×10^-5^ | 0.183 |
| *HMGN1* | protein_coding | HMGN1_1004703954 | CDS | -5.47 | -0.71 | -3.88 | -3.22 | 1.5×10^-4^ | 0.752 |
| *CCDC57* | protein_coding | CCDC57_1001563554 | 3'UTR | -2.92 | -0.71 | -3.50 | -3.49 | 5.9×10^-4^ | 1.000 |
| *DCTN4* | protein_coding | DCTN4_1001667284 | 3'UTR | -3.10 | -0.70 | -3.43 | -3.53 | 7.4×10^-4^ | 1.000 |
| *MYL6* | protein_coding | MYL6_1003679154 | CDS | -5.16 | -0.66 | -3.34 | -3.59 | 0.001 | 1.000 |
| *MBP* | protein_coding | MBP_1004388574 | CDS | -2.44 | 0.67 | 3.22 | -3.67 | 0.002 | 1.000 |
| *TBC1D10B* | protein_coding | TBC1D10B_1001984684 | CDS | -1.92 | 1.03 | 3.21 | -3.67 | 0.002 | 1.000 |
| *CD24* | protein_coding | CD24_1000179484 | 3'UTR | -4.91 | -1.43 | -3.21 | -3.67 | 0.002 | 1.000 |
| *MCM7* | protein_coding | MCM7_1001025304 | CDS | -3.87 | 0.63 | 3.18 | -3.69 | 0.002 | 1.000 |
| *CELSR1* | protein_coding | CELSR1_1001199254 | CDS | -2.77 | -0.61 | -3.16 | -3.70 | 0.002 | 1.000 |
| **Cerebellum (CRB)** |  |  |  |  |  |  |  |  |  |
| *GJA1* | protein_coding | GJA1_1000019024 | CDS | -5.12 | -0.93 | -4.13 | 0.75 | 5.7×10^-5^ | 0.459 |
| *SREK1* | protein_coding | SREK1_1000016244 | CDS | -3.94 | 0.90 | 3.78 | -0.17 | 2.1×10^-4^ | 0.459 |
| *ARGLU1* | protein_coding | ARGLU1_1002109444 | CDS | -4.58 | 1.18 | 3.72 | -0.33 | 2.7×10^-4^ | 0.459 |
| *THRAP3* | protein_coding | THRAP3_1001207614 | CDS | -4.16 | 0.70 | 3.68 | -0.43 | 3.1×10^-4^ | 0.459 |
| *MSANTD4* | protein_coding | MSANTD4_1006252174 | CDS | -3.57 | 0.88 | 3.64 | -0.53 | 3.6×10^-4^ | 0.459 |
| *RIF1* | protein_coding | RIF1_1001776524 | CDS | -3.90 | 1.59 | 3.63 | -0.55 | 3.7×10^-4^ | 0.459 |
| *MORF4L2* | protein_coding | MORF4L2_1006458504 | CDS | -3.97 | 0.71 | 3.61 | -0.60 | 4.0×10^-4^ | 0.459 |
| *NTN4* | protein_coding | NTN4_1006030154 | 3'UTR | -3.96 | 1.46 | 3.59 | -0.66 | 4.3×10^-4^ | 0.459 |
| *THAP12* | protein_coding | THAP12_1001883324 | CDS | -3.22 | -0.90 | -3.57 | -0.69 | 4.5×10^-4^ | 0.459 |
| *MGA* | protein_coding | MGA_1000277994 | CDS | -4.39 | 1.22 | 3.54 | -0.77 | 5.1×10^-4^ | 0.459 |
| **Hippocampus (HIP)** |  |  |  |  |  |  |  |  |  |
| *C12orf49* | protein_coding | C12orf49_1002707404 | 3'UTR | -2.43 | -0.78 | -3.98 | -2.95 | 1.0×10^-4^ | 0.786 |
| *TRIM26* | protein_coding | TRIM26_1002366034 | CDS | -2.17 | -0.72 | -3.87 | -3.04 | 1.5×10^-4^ | 0.786 |
| *TICAM1* | protein_coding | TICAM1_1000957834 | CDS | -2.43 | 0.76 | 3.72 | -3.16 | 2.6×10^-4^ | 0.786 |
| *NR4A2* | protein_coding | NR4A2_1005130524 | CDS | -3.59 | 0.98 | 3.64 | -3.22 | 3.6×10^-4^ | 0.786 |
| *CNOT2* | protein_coding | CNOT2_1004151494 | 3'UTR | -5.36 | 0.67 | 3.55 | -3.29 | 4.9×10^-4^ | 0.786 |
| *ZNFX1* | protein_coding | ZNFX1_1000837204 | 3'UTR | -3.21 | 0.91 | 3.51 | -3.32 | 5.6×10^-4^ | 0.786 |
| *DDX5* | protein_coding | DDX5_1001098634 | CDS | -4.82 | 0.60 | 3.49 | -3.34 | 6.1×10^-4^ | 0.786 |
| *RFX7* | protein_coding | RFX7_1000115374 | CDS | -2.87 | -0.64 | -3.44 | -3.37 | 7.1×10^-4^ | 0.813 |
| *KCTD1* | protein_coding | KCTD1_1006251344 | CDS | -2.34 | -0.80 | -3.34 | -3.45 | 1.0×10^-3^ | 0.857 |
| *ARFGEF3* | protein_coding | ARFGEF3_1004377704 | CDS | -1.95 | -0.90 | -3.34 | -3.45 | 1.0×10^-3^ | 0.857 |
| **Nucleus Accumbens (NAc)** |  |  |  |  |  |  |  |  |  |
| *ZNF740* | protein_coding | ZNF740_1005312874 | 3'UTR | -1.92 | -1.08 | -5.18 | 3.08 | 6.0×10^-7^ | 0.006 |
| *KNOP1* | protein_coding | KNOP1_1005887644 | 3'UTR | -5.03 | 0.94 | 3.77 | -0.60 | 2.2×10^-4^ | 0.567 |
| *ZMYND8* | protein_coding | ZMYND8_1004797514 | CDS | -4.77 | 1.19 | 3.51 | -1.18 | 5.7×10^-4^ | 0.567 |
| *BNC2* | protein_coding | BNC2_1005536874 | CDS | 0.05 | -0.97 | -3.48 | -1.25 | 6.4×10^-4^ | 0.567 |
| *KDELR2* | protein_coding | KDELR2_1001345754 | 3'UTR | -5.96 | 1.06 | 3.45 | -1.30 | 6.9×10^-4^ | 0.567 |
| *ZBTB34* | protein_coding | ZBTB34_1002961124 | CDS | -1.95 | -0.84 | -3.42 | -1.37 | 7.7×10^-4^ | 0.567 |
| *GTPBP2* | protein_coding | GTPBP2_1006111474 | 3'UTR | -2.36 | -0.81 | -3.39 | -1.44 | 8.7×10^-4^ | 0.567 |
| *PAQR4* | protein_coding | PAQR4_1003688114 | 3'UTR | -1.70 | -0.63 | -3.37 | -1.47 | 9.0×10^-4^ | 0.567 |
| *MYEOV* | protein_coding | MYEOV_1002540764 | CDS | -3.05 | -1.13 | -3.37 | -1.49 | 9.3×10^-4^ | 0.567 |
| *BORCS6* | protein_coding | BORCS6_1004678884 | CDS | -1.99 | -0.70 | -3.36 | -1.49 | 9.4×10^-4^ | 0.567 |
| **Prefrontal Cortex (PFC)** |  |  |  |  |  |  |  |  |  |
| *SLC16A1* | protein_coding | SLC16A1_1006255334 | CDS | -2.30 | -1.22 | -5.77 | 7.93 | 3.5×10^-8^ | 3.6×10^-4^ |
| *KNOP1* | protein_coding | KNOP1_1002334624 | 3'UTR | -2.79 | 1.44 | 5.45 | 6.59 | 1.6×10^-7^ | 8.4×10^-4^ |
| *BRPF1* | protein_coding | BRPF1_1002385404 | CDS | -1.92 | 0.74 | 4.92 | 4.44 | 2.0×10^-6^ | 0.006 |
| *MAML2* | protein_coding | MAML2_1004365694 | CDS | -2.22 | -1.14 | -4.87 | 4.28 | 2.4×10^-6^ | 0.006 |
| *DDX39B* | protein_coding | DDX39B_1000001714 | CDS | -1.57 | 0.93 | 4.70 | 3.62 | 5.2×10^-6^ | 0.011 |
| *ZBTB18* | protein_coding | ZBTB18_1002175954 | CDS | -2.60 | -0.89 | -4.63 | 3.35 | 7.1×10^-6^ | 0.012 |
| *NICN1* | protein_coding | NICN1_1005648204 | 3'UTR | -2.31 | 0.74 | 4.44 | 2.67 | 1.6×10^-5^ | 0.023 |
| *SART3* | protein_coding | SART3_1001380654 | 3'UTR | -1.68 | 0.62 | 4.33 | 2.27 | 2.5×10^-5^ | 0.032 |
| *TBC1D13* | protein_coding | TBC1D13_1005655124 | 3'UTR | -2.73 | -0.66 | -4.24 | 1.96 | 3.6×10^-5^ | 0.037 |
| *SH3PXD2B* | protein_coding | SH3PXD2B_1003941164 | 3'UTR | -2.34 | -0.69 | -4.21 | 1.86 | 4.0×10^-5^ | 0.038 |
| **Putamen (PUT)** |  |  |  |  |  |  |  |  |  |
| *LRRN3* | protein_coding | LRRN3_1005814424 | CDS | -2.51 | -0.78 | -4.17 | -2.64 | 4.8×10^-5^ | 0.491 |
| *ZNF264* | protein_coding | ZNF264_1005703964 | CDS | -2.07 | 0.79 | 3.88 | -2.91 | 1.5×10^-4^ | 0.666 |
| *PLK2* | protein_coding | PLK2_1004342404 | 3'UTR | -4.46 | 0.77 | 3.76 | -3.01 | 2.3×10^-4^ | 0.666 |
| *TACC3* | protein_coding | TACC3_1004946464 | CDS | -2.90 | -0.73 | -3.58 | -3.16 | 4.5×10^-4^ | 0.666 |
| *LOC642852* | ncRNA | LOC642852_1004175924 | 283 | -3.82 | -0.71 | -3.56 | -3.17 | 4.7×10^-4^ | 0.666 |
| *CDC42EP4* | protein_coding | CDC42EP4_1002504694 | CDS | -2.16 | -0.88 | -3.56 | -3.18 | 4.8×10^-4^ | 0.666 |
| *CARM1* | protein_coding | CARM1_1005728104 | CDS | -2.43 | 0.77 | 3.54 | -3.19 | 5.1×10^-4^ | 0.666 |
| *ATF7* | protein_coding | ATF7_1004207914 | 3'UTR | -2.49 | 0.96 | 3.50 | -3.23 | 6.0×10^-4^ | 0.666 |
| *PLK2* | protein_coding | PLK2_1004979524 | 3'UTR | -4.44 | 0.83 | 3.45 | -3.27 | 7.0×10^-4^ | 0.666 |
| *STIMATE* | protein_coding | STIMATE_1003300074 | 3'UTR | -2.32 | 0.62 | 3.45 | -3.27 | 7.0×10^-4^ | 0.666 |
| **Ventral Tegmental Area (VTA)** |  |  |  |  |  |  |  |  |  |
| *CIC* | protein_coding | CIC_1001657134 | 3'UTR | -2.05 | 0.77 | 4.74 | 3.19 | 4.3×10^-6^ | 0.044 |
| *RBM15* | protein_coding | RBM15_1005954584 | CDS | -2.82 | -0.68 | -4.29 | 1.70 | 2.9×10^-5^ | 0.151 |
| *ANKH* | protein_coding | ANKH_1006190994 | 3'UTR | -4.23 | 0.74 | 4.19 | 1.39 | 4.4×10^-5^ | 0.151 |
| *ERICH1* | protein_coding | ERICH1_1004319574 | CDS | -1.40 | 0.80 | 4.08 | 1.07 | 6.6×10^-5^ | 0.170 |
| *MAN1B1* | protein_coding | MAN1B1_1005550234 | 3'UTR | -3.20 | 0.67 | 3.86 | 0.42 | 1.6×10^-4^ | 0.250 |
| *PTPDC1* | protein_coding | PTPDC1_1002284614 | CDS | -2.29 | 0.69 | 3.85 | 0.39 | 1.6×10^-4^ | 0.250 |
| *TET2* | protein_coding | TET2_1004935904 | CDS | -2.14 | -0.59 | -3.84 | 0.35 | 1.7×10^-4^ | 0.250 |
| *SH3PXD2A* | protein_coding | SH3PXD2A_1005271274 | CDS | -2.24 | 0.68 | 3.76 | 0.13 | 2.3×10^-4^ | 0.292 |
| *PAQR4* | protein_coding | PAQR4_1000321814 | 3'UTR | -5.45 | 0.76 | 3.67 | -0.13 | 3.2×10^-4^ | 0.325 |
| *MTCL1* | protein_coding | MTCL1_1005694634 | CDS | -1.98 | -0.85 | -3.61 | -0.30 | 4.0×10^-4^ | 0.325 |

Probe Name: Gene name_m_6_A probe ID.

Log_2_FC: the log_2_ fold-change (FC) between cases and controls.

AveMeth: the average methylation level of m^6^A sites.

t: the t-statistics used to assess differential methylation.

B: the empirical Bayes log odds of differential methylation.

*P*-value: the *P*-value for differential methylation (unadjusted for multiple testing).

*P*_adj_: the *P*-value adjusted for multiple testing (the default is Benjamini-Hochberg).
